# Lipid peroxidation and colorectal cancer risk: a time-varying relationship

**DOI:** 10.1101/2025.02.16.25322362

**Authors:** Gong Yang, Ginger L. Milne, Marina S. Nogueira, Haoyang Yi, Qing Lan, Yu-Tang Gao, Xiao-Ou Shu, Wei Zheng, Qingxia Chen

## Abstract

**Importance:** We recently observed an inverse and time-dependent association between systemic oxidative stress (OxS), measured by urinary biomarkers of nucleic acid oxidation, and colorectal cancer (CRC) risk. Further investigations into other types of OxS markers are warranted.

**Objective:** To extend the investigation into systemic lipid peroxidation and CRC risk.

**Design, Setting, and Participants:** Utilizing a nested case-control design, this study’s primary analysis was performed in two large prospective cohorts in Shanghai, China, and replicated in an independent cohort in the US.

**Exposures:** Systemic lipid peroxidation was assessed by urinary F_2_-isoprostanes (F_2_-IsoPs) using UPLC-MS/MS assays.

**Main Outcomes and Measures:** During 15.1-year follow-up in the Shanghai cohorts, 1938 incident CRC cases were identified and matched to one control each through incidence-density sampling. In the US cohort, 285 incident CRC cases were included, each matched to two controls. Odds ratios (ORs) for CRC were calculated using multivariable conditional logistic regression models.

**Results:** Elevated levels of urinary 5-F_2t_-IsoP (5-iPF_2α_-VI), a major isomer of F_2_-IsoPs induced solely by free radicals, were associated with reduced risk of CRC in the Shanghai cohorts. This finding was replicated in the US cohort. Moreover, this inverse association was time-dependent, manifesting only in the later years of cancer development. Multivariable-adjusted ORs (95% CI) for CRC diagnosed within 5 years of enrollment at the 10th and 90th percentiles of 5-F_2t_-IsoP levels, relative to the median, were 1.57 (1.26-1.96) and 0.61 (0.42-0.89), respectively, indicating a 2.2-fold difference in risk between the two groups. A stronger association was observed when using the composite index of DNA, RNA and lipid OxS markers, showing a 3.9-fold difference in risk between the two groups. No significant association was found for CRC diagnosed beyond 5 years of enrollment.

**Conclusions:** This study provides new evidence that systemic OxS is inversely and time-dependently associated with CRC risk in humans.

**Key Points:** *Question:* Is the time-dependent relationship between oxidative stress and tumorigenesis observed at the cellular level in experimental models also present at the systemic level in a population-based setting?

*Findings:* Elevated systemic lipid peroxidation, measured by urinary F_2_-isoprostanes, was associated with a reduced risk of colorectal cancer (CRC) in two large prospective cohort studies in Shanghai, China, which was replicated in an independent cohort in the United States. This association varied over time, showing a stronger effect as cancer advanced.

*Meaning:* This study provides new evidence that systemic oxidative stress is inversely and time-dependently associated with CRC risk in humans.

## INTRODUCTION

We recently observed that elevated systemic oxidative stress (OxS), measured by DNA (8-oxo-dG) and RNA oxidation biomarkers (8-oxo-Guo) in urine, was associated with reduced risk of colorectal cancer (CRC) in three large prospective cohorts.^1^ This relationship was time-dependent, becoming stronger as the OxS assessment approaches the time of CRC diagnosis. This finding provides the first evidence that the time-dependent relationship between OxS and tumorigenesis, previously observed at the cellular level in experimental models,^2–4^ also exists at the systemic level in a population-based setting. Additionally, this finding offers an alternative mechanistic insight into the long-standing question of why antioxidant-based chemoprevention has led to detrimental outcomes among high-risk individuals in large randomized controlled trials (RCTs).^5–7^

Importantly, levels of nucleic acid oxidation biomarkers reflect both free radical-induced formation and enzymatic repair of oxidative damage.^8^ Thus, the potential influence of DNA repair capability on OxS assessment could complicate or obscure causal inference, making it difficult to attribute the study finding solely to free radical-induced damage. Moreover, diverse effects of oxidative damage to various macromolecules from different cellular compartments (e.g. membrane lipids, nucleus DNA, and cytoplasmic RNA) on tumorigenesis have been suggested.^9^ In this report, we extended our study to examine whether the time-dependent association with DNA and RNA OxS markers can be replicated in lipid peroxidation markers, and whether an index of three types (DNA, RNA, and lipid) of OxS markers can better assess the relationship with CRC risk in the same study populations.

## METHODS

### Study Population

The present study leverages two large population-based cohorts in Shanghai, China―the Shanghai Women’s Health Study (SWHS) and the Shanghai Men’s Health Study (SMHS)―for the primary analysis,^10,11^ and an independent cohort study in the United States―the Southern Community Cohort Study (SCCS)―for replication.^12^ Details of the design and methods for the SWHS and SMHS have been reported elsewhere.^10,11^ Briefly, 74 941 females aged 40-70 years were recruited to the SWHS between 1996 and 2000, while 61 480 males aged 40-74 years were recruited to the SMHS between 2002 and 2006. The SCCS is a prospective cohort study conducted across 12 southern US states, enrolling over 85 000 participants aged 40-79 years between 2002 and 2009.^12^ Baseline exposures were assessed through in-person interviews in the three cohorts, and spot urine samples were collected from 87.7% SWHS, 89.1% SMHS, and 27.6% SCCS participants, after obtaining written informed consent. Protocols for the three cohort studies were approved by institutional review boards for human research at all participating institutes.

### Follow-up and cancer identification

Cohort members in the SWHS and SMHS have been followed for occurrence of cancer through a combination of repeated home visits and annual record linkage to Shanghai Tumor Registry and Shanghai Vital Statistics. The diagnosis was verified via reviewing medical records. Incident cancer cases in the SCCS were ascertained through linkage to 12 US state cancer registries and National Death Index mortality records. CRC cases included in this study were cancer-free at baseline and were diagnosed with CRC as the first primary cancer during follow-up.

### The nested case-control study

Utilizing a nested case-control design, this study’s primary analysis was performed in the SWHS and SMSH and replicated in the SCCS. During a median follow-up of 15.1 years in the SWHS and SMHS, 1938 incident cases of CRC (1035 females and 903 males) with baseline urine samples were identified and included in this study. For each case, one control was randomly selected using incidence-density sampling to individually match on age at baseline (±2 years), sex, date (±30 days) and time (morning/afternoon) of sample collection, and interval since the last meal (±2 hours) among those who provided baseline urine samples and were cancer-free at the time of cancer diagnosis for the index case. In the SCCS, 285 incident CRC cases were identified among cancer-free participants with baseline urine samples. Each case was matched to two controls using the same criteria from the Shanghai cohorts, plus race. Non-Black or nor-White participants (4.9%) were not included due to concerns on low statistical power in those groups.

### Laboratory Measurements

Selected F_2_-IsoP biomarkers were measured with liquid chromatography-mass spectrometry (UPLC-MS/MS) by the VUMC Eicosanoid Core Laboratory (**Supplementary Method**). Sixty-four F_2_-IsoP isomers are generated from the free radical peroxidation of arachidonic acid. In this study, the isomers 5-F_2t_-IsoP, 15-F_2t_-IsoP and its β-oxidation product 2,3-dinor-5,6-dihydro-15-F_2t_-IsoP (15-F_2t_-IsoP-M) were quantified using commercially available standards (Cayman Chemical Company, Ann Arbor, MI USA). The 5– and 15-series F_2_-IsoPs are known to be formed in the greatest abundance.^13,14^ Samples from each case-control pair were analyzed in the same assay batch with a random and blind arrangement. The average within-day and between-day assay coefficients of variation (CV) are typically lower than 8%. Outliers with intra-batch CV >20% were excluded from the main analysis. All measurements were standardized for variations in urinary flow using urine creatinine and expressed as nanograms per milligram of creatinine (ng/mg Cr).

### Statistical Analysis

The primary analysis was conducted in the SWHS and SMHS and followed by an independent replication in the SCCS. Conditional logistic regression models, with matched case-control sets as strata, were used to estimate odds ratios (ORs) and 95% confidence intervals (CIs) for CRC risk in association with selected biomarkers.

Restricted cubic spline functions with 3 knots were applied to evaluate potential nonlinear effects,^15^ and the exposure and outcome relationship was graphically illustrated. Exposures were categorized based on percentile distributions in controls, with ORs presented for the 10th, 30th, 70th, and 90th percentiles, compared to the 50th percentile (reference).

Potential confounders adjusted for in the multivariable model included age, education, cigarette smoking, alcohol consumption, body mass index (BMI), physical activity, menopausal status (for female participants), regular use of vitamin supplements, regular use of aspirin and other nonsteroidal anti-inflammatory drugs (NSAIDs), Charlson comorbidity score, history of CRC in first-degree relatives, and total energy intake. Stratified analyses by time elapsed from baseline to cancer diagnosis were performed to examine potential time-varying relationships.

Comprehensive approaches to handling intra– and inter-batch variations were detailed in the **Supplementary Method**. Biomarker measures from batches with intra-batch CV <20% were included in the main analysis, encompassing 83.4% of study samples for 15-F_2t_-IsoP, 91.7% for 5-F_2t_-IsoP, and 99.0% for 15-F_2t_-IsoP-M in the SWHS and SMHS, and 100% of study samples for all selected markers in the SCCS. Inter-batch effects were corrected by centering marker measurements using the median value of QC samples within each batch.^16^ As samples from each case-control pair were assayed in the same batch, comparisons between cases and controls were expected to be robust to the inter-batch normalization process. Nonetheless, sensitivity analyses were conducted on the assay data without inter-batch effect correction in two scenarios: 1) analyses including samples with intra-batch CV <20%, and 2) analyses involving all samples regardless of their intra-batch CVs.

A composite index was constructed based on the principal component analysis of three OxS markers: DNA (8-oxo-dG), RNA (8-oxo-Guo), and lipid markers (5-F_2t_-IsoP). The index is formulated: 0.61*8-Oxo-dG+0.62*8-Oxo-Guo+0.49*5-F_2t_-IsoP. All statistical analyses were conducted using R software (version 4.0.4) with significance defined as two-sided *P* < .05.

## RESULTS

### Characteristics of study participants

The primary analysis included 1938 CRC case-control pairs from the SWHS and SMHS, with a mean age (SD) at diagnosis of 68.3 (9.3) years, and 53.4% were female. Cases and controls were largely comparable in demographics, lifestyle, and known risk factors, except for a higher likelihood of CRC family history in cases (**Supplementary Table 1**).

In the replication cohort SCCS, 285 CRC cases and 570 matched controls were included. The mean age (SD) at CRC diagnosis was 55.1 (8.8) years, with 67.4% African Americans and 33.6% European Americans. Baseline characteristics were generally comparable between case and control groups (**Supplementary Table 2**).

### Association of CRC risk with urinary 5-F_2t_-IsoP

In the SWHS and SMHS, levels of urinary 5-F_2t_-IsoP, formed solely via free radical lipid peroxidation without influence from the cyclooxygenases, were significantly lower in CRC cases than in controls **(Supplementary Table 3).** The geometric mean (95% CI) was 6.22 ng/mg Cr (6.02-6.43) for CRC cases compared to 6.76 ng/mg Cr (6.54-6.99) for controls (*P* < .001). A similar case-control difference was suggested in the SCCS, although statistically insignificant.

**Table 1** shows that elevated 5-F_2t_-IsoP levels at baseline were significantly associated with reduced subsequent risk of CRC in the SWHS and SMHS (overall *P* =.001). The multivariable-adjusted ORs (95% CI) for CRC at the 10th and 90th percentiles of 5-F_2t_-IsoP levels, relative to the median, were 1.12 (1.04-1.21) and 0.84 (0.73-0.97), respectively. This inversion association was replicated in the SCCS (overall *P* = .02), with corresponding ORs (95% CI) of 1.38 (1.08-1.76) and 0.84 (0.67-1.06), respectively.

**Table 1.**
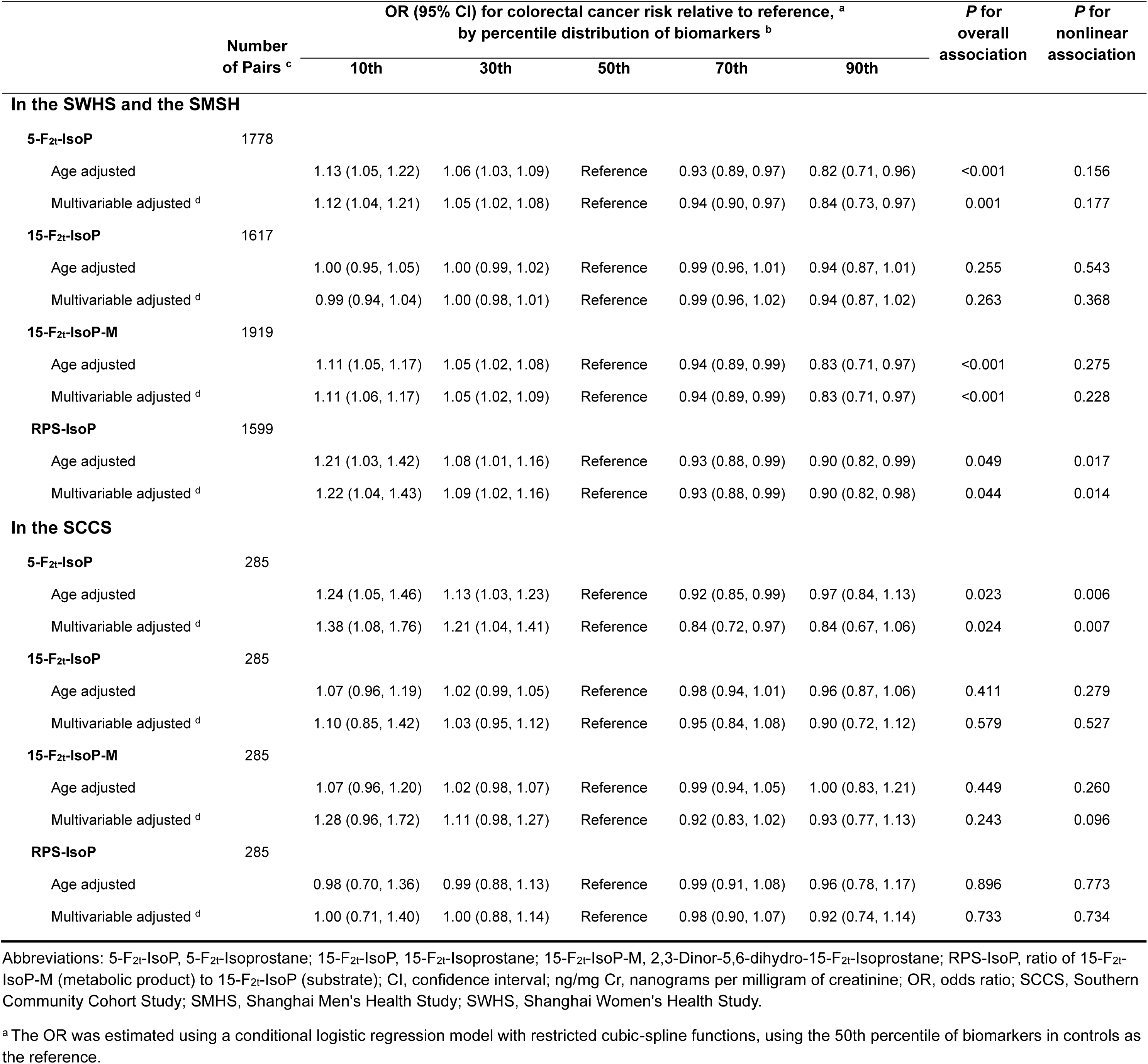
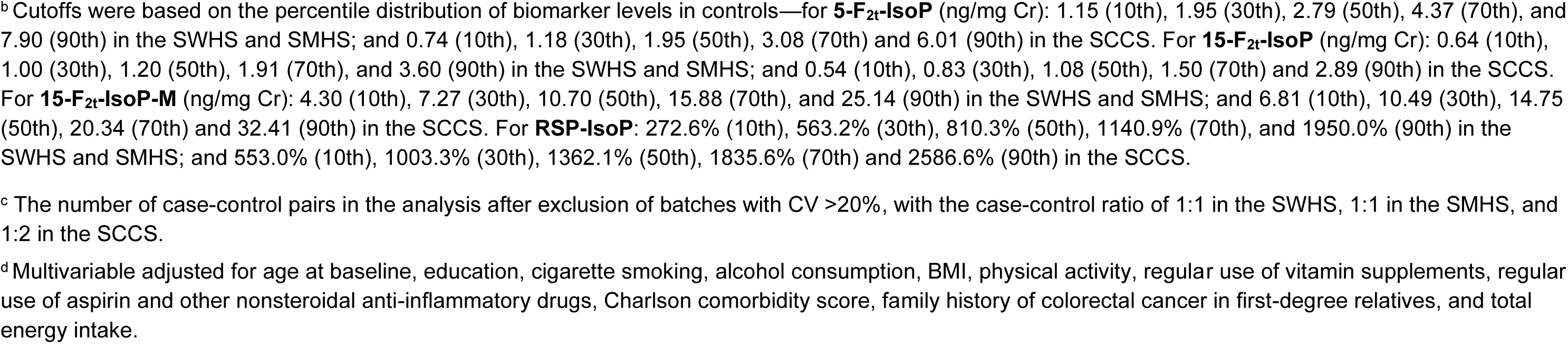
Associations between baseline urinary levels of F_2_-IsoPs and subsequent risk of colorectal cancer.

|  | Number<br>of Pairs <sup>c</sup> | OR (95% CI) for colorectal cancer risk relative to reference, <sup>a</sup><br>by percentile distribution of biomarkers <sup>b</sup> |  |  |  |  | *P* for<br>overall<br>association | *P* for<br>nonlinear<br>association |
| --- | --- | --- | --- | --- | --- | --- | --- | --- |
|  |  | 10th | 30th | 50th | 70th | 90th |  |  |
| In the SWHS and the SMSH |  |  |  |  |  |  |  |  |
| 5-F<sub>2t</sub>-IsoP | 1778 |  |  |  |  |  |  |  |
| Age adjusted |  | 1.13 (1.05, 1.22) | 1.06 (1.03, 1.09) | Reference | 0.93 (0.89, 0.97) | 0.82 (0.71, 0.96) | <0.001 | 0.156 |
| Multivariable adjusted <sup>d</sup> |  | 1.12 (1.04, 1.21) | 1.05 (1.02, 1.08) | Reference | 0.94 (0.90, 0.97) | 0.84 (0.73, 0.97) | 0.001 | 0.177 |
| 15-F<sub>2t</sub>-IsoP | 1617 |  |  |  |  |  |  |  |
| Age adjusted |  | 1.00 (0.95, 1.05) | 1.00 (0.99, 1.02) | Reference | 0.99 (0.96, 1.01) | 0.94 (0.87, 1.01) | 0.255 | 0.543 |
| Multivariable adjusted <sup>d</sup> |  | 0.99 (0.94, 1.04) | 1.00 (0.98, 1.01) | Reference | 0.99 (0.96, 1.02) | 0.94 (0.87, 1.02) | 0.263 | 0.368 |
| 15-F<sub>2t</sub>-IsoP-M | 1919 |  |  |  |  |  |  |  |
| Age adjusted |  | 1.11 (1.05, 1.17) | 1.05 (1.02, 1.08) | Reference | 0.94 (0.89, 0.99) | 0.83 (0.71, 0.97) | <0.001 | 0.275 |
| Multivariable adjusted <sup>d</sup> |  | 1.11 (1.06, 1.17) | 1.05 (1.02, 1.09) | Reference | 0.94 (0.89, 0.99) | 0.83 (0.71, 0.97) | <0.001 | 0.228 |
| RPS-IsoP | 1599 |  |  |  |  |  |  |  |
| Age adjusted |  | 1.21 (1.03, 1.42) | 1.08 (1.01, 1.16) | Reference | 0.93 (0.88, 0.99) | 0.90 (0.82, 0.99) | 0.049 | 0.017 |
| Multivariable adjusted <sup>d</sup> |  | 1.22 (1.04, 1.43) | 1.09 (1.02, 1.16) | Reference | 0.93 (0.88, 0.99) | 0.90 (0.82, 0.98) | 0.044 | 0.014 |
| In the SCCS |  |  |  |  |  |  |  |  |
| 5-F<sub>2t</sub>-IsoP | 285 |  |  |  |  |  |  |  |
| Age adjusted |  | 1.24 (1.05, 1.46) | 1.13 (1.03, 1.23) | Reference | 0.92 (0.85, 0.99) | 0.97 (0.84, 1.13) | 0.023 | 0.006 |
| Multivariable adjusted <sup>d</sup> |  | 1.38 (1.08, 1.76) | 1.21 (1.04, 1.41) | Reference | 0.84 (0.72, 0.97) | 0.84 (0.67, 1.06) | 0.024 | 0.007 |
| 15-F<sub>2t</sub>-IsoP | 285 |  |  |  |  |  |  |  |
| Age adjusted |  | 1.07 (0.96, 1.19) | 1.02 (0.99, 1.05) | Reference | 0.98 (0.94, 1.01) | 0.96 (0.87, 1.06) | 0.411 | 0.279 |
| Multivariable adjusted <sup>d</sup> |  | 1.10 (0.85, 1.42) | 1.03 (0.95, 1.12) | Reference | 0.95 (0.84, 1.08) | 0.90 (0.72, 1.12) | 0.579 | 0.527 |
| 15-F<sub>2t</sub>-IsoP-M | 285 |  |  |  |  |  |  |  |
| Age adjusted |  | 1.07 (0.96, 1.20) | 1.02 (0.98, 1.07) | Reference | 0.99 (0.94, 1.05) | 1.00 (0.83, 1.21) | 0.449 | 0.260 |
| Multivariable adjusted <sup>d</sup> |  | 1.28 (0.96, 1.72) | 1.11 (0.98, 1.27) | Reference | 0.92 (0.83, 1.02) | 0.93 (0.77, 1.13) | 0.243 | 0.096 |
| RPS-IsoP | 285 |  |  |  |  |  |  |  |
| Age adjusted |  | 0.98 (0.70, 1.36) | 0.99 (0.88, 1.13) | Reference | 0.99 (0.91, 1.08) | 0.96 (0.78, 1.17) | 0.896 | 0.773 |
| Multivariable adjusted <sup>d</sup> |  | 1.00 (0.71, 1.40) | 1.00 (0.88, 1.14) | Reference | 0.98 (0.90, 1.07) | 0.92 (0.74, 1.14) | 0.733 | 0.734 |

Abbreviations: 5-F<sub>2t</sub>-IsoP, 5-F<sub>2t</sub>-Isoprostane; 15-F<sub>2t</sub>-IsoP, 15-F<sub>2t</sub>-Isoprostane; 15-F<sub>2t</sub>-IsoP-M, 2,3-Dinor-5,6-dihydro-15-F<sub>2t</sub>-Isoprostane; RPS-IsoP, ratio of 15-F<sub>2t</sub>-IsoP-M (metabolic product) to 15-F<sub>2t</sub>-IsoP (substrate); CI, confidence interval; ng/mg Cr, nanograms per milligram of creatinine; OR, odds ratio; SCCS, Southern Community Cohort Study; SMHS, Shanghai Men's Health Study; SWHS, Shanghai Women's Health Study.
<sup>a</sup> The OR was estimated using a conditional logistic regression model with restricted cubic-spline functions, using the 50th percentile of biomarkers in controls as the reference.
<sup>b</sup> Cutoffs were based on the percentile distribution of biomarker levels in controls—for **5-F<sub>2t</sub>-IsoP** (ng/mg Cr): 1.15 (10th), 1.95 (30th), 2.79 (50th), 4.37 (70th), and 7.90 (90th) in the SWHS and SMHS; and 0.74 (10th), 1.18 (30th), 1.95 (50th), 3.08 (70th) and 6.01 (90th) in the SCCS. For **15-F<sub>2t</sub>-IsoP** (ng/mg Cr): 0.64 (10th), 1.00 (30th), 1.20 (50th), 1.91 (70th), and 3.60 (90th) in the SWHS and SMHS; and 0.54 (10th), 0.83 (30th), 1.08 (50th), 1.50 (70th) and 2.89 (90th) in the SCCS. For **15-F<sub>2t</sub>-IsoP-M** (ng/mg Cr): 4.30 (10th), 7.27 (30th), 10.70 (50th), 15.88 (70th), and 25.14 (90th) in the SWHS and SMHS; and 6.81 (10th), 10.49 (30th), 14.75 (50th), 20.34 (70th) and 32.41 (90th) in the SCCS. For **RSP-IsoP**: 272.6% (10th), 563.2% (30th), 810.3% (50th), 1140.9% (70th), and 1950.0% (90th) in the SWHS and SMHS; and 553.0% (10th), 1003.3% (30th), 1362.1% (50th), 1835.6% (70th) and 2586.6% (90th) in the SCCS.
<sup>c</sup> The number of case-control pairs in the analysis after exclusion of batches with CV >20%, with the case-control ratio of 1:1 in the SWHS, 1:1 in the SMHS, and 1:2 in the SCCS.
<sup>d</sup> Multivariable adjusted for age at baseline, education, cigarette smoking, alcohol consumption, BMI, physical activity, regular use of vitamin supplements, regular use of aspirin and other nonsteroidal anti-inflammatory drugs, Charlson comorbidity score, family history of colorectal cancer in first-degree relatives, and total energy intake.

We further analyzed if the observed inverse association was time-dependent, given a time-varying relationship with DNA/RNA OxS markers revealed in our earlier study.^1^ Due to the small number of cases in the SCCS (n=285), analyses stratified by follow-up time were performed only in the Shanghai cohorts (n=1938). CRC cases were categorized based on the time from enrollment to CRC diagnosis: within 5 years, between 5 and 9 years, and beyond 9 years. Stratified analyses revealed a pronounced temporal pattern in the 5-F_2t_-IsoP and CRC association, with *P* for heterogeneity of < .001 (**Figure 1**). Urinary 5-F_2t_-IsoP levels were strongly and inversely associated with CRC risk diagnosed within 5 years of enrollment (overall *P* < .0001). ORs (95% CI) for CRC at the 10th and 90th percentiles of 5-F_2t_-IsoP, relative to the median, were 1.57 (1.26-1.96) and 0.61 (0.42-0.89), respectively, demonstrating a 2.2-fold difference in risk between the two groups (**Table 2**). However, no significant association was found for CRC diagnosed beyond 5 years of enrollment.

**Figure 1.**
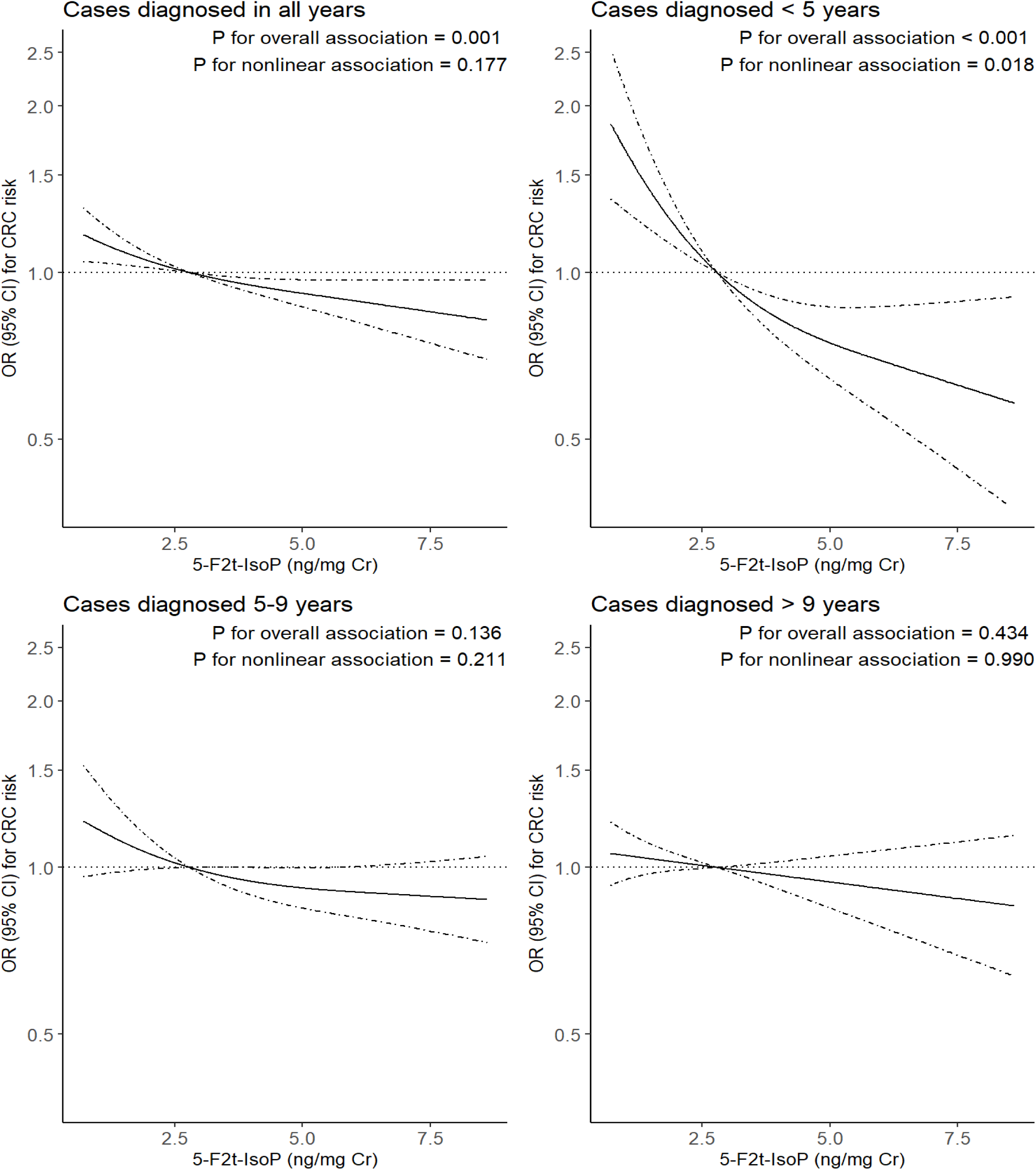
Smoothed plot for multivariable-adjusted odds ratios for colorectal cancer risk according to urinary levels of 5-F_2t_-IsoP by time interval from baseline to cancer diagnosis: Overall, <5 years, 5-9 years and >9 years in the SWHS and the SMHS. The OR was estimated using a conditional logistic regression model with restricted cubic spline functions, with the 50th percentile of biomarker levels in controls as the reference and adjusted for potential confounding factors as listed in the footnotes of Table 1. OR values on the *y* axis are shown in log scale. The solid line indicates the point estimate, and the dashed lines indicate the 95% CI. Abbreviations: 5-F_2t_-IsoP, 5-F_2t_-Isoprostane; CI, confidence interval; ng/mg Cr, nanograms per milligram of creatinine; OR, odds ratio; SMHS, Shanghai Men’s Health Study; SWHS, Shanghai Women’s Health Study.

**Table 2.** Associations between baseline urinary levels of F_2_-IsoPs and subsequent risk of colorectal cancer by time interval between urine sample collection at baseline and cancer diagnosis during follow-up in the SWHS and SMHS.

|  |  | OR (95% CI) for colorectal cancer risk relative to reference, <sup>a</sup><br>by percentile distribution of biomarkers <sup>b</sup> |  |  |  |  | *P* for<br>overall<br>association | *P* for<br>nonlinear<br>association |
| --- | --- | --- | --- | --- | --- | --- | --- | --- |
|  | Number<br>of Pairs <sup>c</sup> | 10th | 30th | 50th | 70th | 90th |  |  |
| Cases diagnosed <5 years after enrollment |  |  |  |  |  |  |  |  |
| 5-F<sub>2t</sub>-IsoP | 387 |  |  |  |  |  |  |  |
| Age adjusted |  | 1.51 (1.22, 1.86) | 1.20 (1.10, 1.31) | Reference | 0.80 (0.72, 0.89) | 0.61 (0.42, 0.89) | <0.001 | 0.036 |
| Multivariable adjusted <sup>d</sup> |  | 1.57 (1.26, 1.96) | 1.22 (1.11, 1.33) | Reference | 0.79 (0.71, 0.88) | 0.61 (0.42, 0.89) | <0.001 | 0.018 |
| 15-F<sub>2t</sub>-IsoP | 367 |  |  |  |  |  |  |  |
| Age adjusted |  | 1.06 (0.96, 1.18) | 1.02 (0.99, 1.05) | Reference | 0.96 (0.90, 1.03) | 0.95 (0.82, 1.10) | 0.522 | 0.311 |
| Multivariable adjusted <sup>d</sup> |  | 1.07 (0.96, 1.20) | 1.02 (0.99, 1.05) | Reference | 0.96 (0.90, 1.03) | 0.96 (0.83, 1.12) | 0.477 | 0.238 |
| 15-F<sub>2t</sub>-IsoP-M | 411 |  |  |  |  |  |  |  |
| Age adjusted |  | 1.09 (0.97, 1.23) | 1.03 (0.96, 1.10) | Reference | 0.98 (0.87, 1.11) | 0.96 (0.68, 1.35) | 0.093 | 0.129 |
| Multivariable adjusted <sup>d</sup> |  | 1.08 (0.96, 1.23) | 1.03 (0.95, 1.10) | Reference | 0.99 (0.87, 1.12) | 0.96 (0.67, 1.38) | 0.137 | 0.145 |
| RPS-IsoP | 349 |  |  |  |  |  |  |  |
| Age adjusted |  | 1.04 (0.74, 1.46) | 1.02 (0.89, 1.16) | Reference | 0.99 (0.89, 1.10) | 0.99 (0.84, 1.16) | 0.747 | 0.798 |
| Multivariable adjusted <sup>d</sup> |  | 1.04 (0.73, 1.47) | 1.01 (0.88, 1.16) | Reference | 0.99 (0.89, 1.11) | 0.99 (0.84, 1.17) | 0.731 | 0.829 |
| Cases diagnosed 5-9 years after enrollment |  |  |  |  |  |  |  |  |
| 5-F<sub>2t</sub>-IsoP | 469 |  |  |  |  |  |  |  |
| Age adjusted |  | 1.18 (1.01, 1.39) | 1.08 (1.01, 1.15) | Reference | 0.92 (0.86, 0.98) | 0.87 (0.75, 1.01) | 0.054 | 0.116 |
| Multivariable adjusted <sup>d</sup> |  | 1.15 (0.98, 1.36) | 1.06 (0.99, 1.14) | Reference | 0.93 (0.87, 1.00) | 0.88 (0.75, 1.03) | 0.136 | 0.211 |
| 15-F<sub>2t</sub>-IsoP | 358 |  |  |  |  |  |  |  |
| Age adjusted |  | 1.03 (0.92, 1.15) | 1.01 (0.98, 1.04) | Reference | 0.97 (0.91, 1.03) | 0.91 (0.79, 1.03) | 0.325 | 0.955 |
| Multivariable adjusted <sup>d</sup> |  | 1.01 (0.89, 1.14) | 1.00 (0.97, 1.04) | Reference | 0.98 (0.92, 1.05) | 0.93 (0.82, 1.06) | 0.529 | 0.849 |
| 15-F<sub>2t</sub>-IsoP-M | 539 |  |  |  |  |  |  |  |
| Age adjusted |  | 1.10 (1.02, 1.20) | 1.05 (1.00, 1.09) | Reference | 0.94 (0.87, 1.02) | 0.85 (0.68, 1.07) | 0.058 | 0.450 |
| Multivariable adjusted <sup>d</sup> |  | 1.11 (1.02, 1.21) | 1.05 (1.00, 1.10) | Reference | 0.94 (0.87, 1.02) | 0.85 (0.68, 1.07) | 0.041 | 0.333 |
| RPS-IsoP | 358 |  |  |  |  |  |  |  |
| Age adjusted |  | 1.17 (0.85, 1.62) | 1.07 (0.93, 1.22) | Reference | 0.95 (0.85, 1.06) | 0.92 (0.78, 1.09) | 0.614 | 0.337 |
| Multivariable adjusted <sup>d</sup> |  | 1.27 (0.90, 1.78) | 1.10 (0.96, 1.27) | Reference | 0.92 (0.82, 1.04) | 0.89 (0.74, 1.06) | 0.382 | 0.171 |
| Cases diagnosed >9 years after enrollment |  |  |  |  |  |  |  |  |
| 5-F<sub>2t</sub>-IsoP | 922 |  |  |  |  |  |  |  |
| Age adjusted |  | 1.06 (0.98, 1.16) | 1.03 (1.00, 1.07) | Reference | 0.95 (0.88, 1.02) | 0.84 (0.65, 1.08) | 0.231 | 0.914 |
| Multivariable adjusted <sup>d</sup> |  | 1.05 (0.96, 1.14) | 1.02 (0.99, 1.06) | Reference | 0.96 (0.89, 1.03) | 0.87 (0.67, 1.12) | 0.434 | 0.990 |
| **15-F<sub>2t</sub>-IsoP** | 892 |  |  |  |  |  |  |  |
| Age adjusted |  | 0.96 (0.89, 1.03) | 0.99 (0.98, 1.01) | Reference | 0.98 (0.94, 1.03) | 0.90 (0.76, 1.06) | 0.196 | 0.095 |
| Multivariable adjusted <sup>d</sup> |  | 0.96 (0.89, 1.03) | 0.99 (0.97, 1.01) | Reference | 0.98 (0.94, 1.03) | 0.91 (0.77, 1.08) | 0.242 | 0.107 |
| **15-F<sub>2t</sub>-IsoP-M** | 969 |  |  |  |  |  |  |  |
| Age adjusted |  | 1.14 (1.05, 1.24) | 1.07 (1.02, 1.13) | Reference | 0.90 (0.82, 0.99) | 0.74 (0.57, 0.97) | 0.008 | 0.998 |
| Multivariable adjusted <sup>d</sup> |  | 1.14 (1.04, 1.24) | 1.07 (1.02, 1.13) | Reference | 0.90 (0.82, 0.99) | 0.75 (0.57, 0.98) | 0.013 | 0.983 |
| **RPS-IsoP** | 892 |  |  |  |  |  |  |  |
| Age adjusted |  | 1.31 (1.06, 1.62) | 1.13 (1.03, 1.24) | Reference | 0.89 (0.81, 0.98) | 0.84 (0.72, 0.98) | 0.036 | 0.010 |
| Multivariable adjusted <sup>d</sup> |  | 1.31 (1.05, 1.62) | 1.12 (1.02, 1.24) | Reference | 0.89 (0.81, 0.98) | 0.84 (0.72, 0.98) | 0.046 | 0.013 |

Abbreviations: 5-F<sub>2t</sub>-IsoP, 5-F<sub>2t</sub>-Isoprostane; 15-F<sub>2t</sub>-IsoP, 15-F<sub>2t</sub>-Isoprostane; 15-F<sub>2t</sub>-IsoP-M, 2,3-Dinor-5,6-dihydro-15-F<sub>2t</sub>-Isoprostane; RPS-IsoP, ratio of 15-F<sub>2t</sub>-IsoP-M (metabolic product) to 15-F<sub>2t</sub>-IsoP (substrate); CI, confidence interval; ng/mg Cr, nanograms per milligram of creatinine; OR, odds ratio; SMHS, Shanghai Men's Health Study; SWHS, Shanghai Women's Health Study.
<sup>a</sup> The OR was estimated using a conditional logistic regression model with restricted cubic-spline functions, using the 50th percentile of biomarkers in controls as the reference.
<sup>b</sup> Cutoffs were based on the percentile distribution of biomarker levels in controls as detailed in the footnotes of Table 1.
<sup>c</sup> The number of case-control pairs in the analysis after exclusion of batches with CV >20%.
<sup>d</sup> Multivariable adjusted for age at baseline, education, cigarette smoking, alcohol consumption, BMI, physical activity, regular use of vitamin supplements, regular use of aspirin and other nonsteroidal anti-inflammatory drugs, Charlson comorbidity score, family history of colorectal cancer in first-degree relatives, and total energy intake.

### Association of CRC risk with urinary 15-F_2t_-IsoP

Urinary 15-F_2t_-IsoP (also known as 8-iso-PGF_2a_), the most intensively studied biomarker of lipid peroxidation, can be generated nonenzymatically by lipid peroxidation and enzymatically by cyclooxygenase enzymes.^17^ In both the primary and replication cohorts, there were no significant differences in 15-F_2t_-IsoP levels between CRC cases and controls **(Supplementary Table 3)**, resulting in a null association for overall CRC risk (**Table 1**) or CRC risk across different follow-up windows (**Table 2** and **Supplementary** Figure 1).

### Association of CRC risk with β-oxidation metabolite of 15-F_2t_-IsoP

15-F_2t_-IsoP is metabolized to 15-F_2t_-IsoP-M via β-oxidation. Levels of 15-F_2t_-IsoP-M, unlike unmetabolized 15-F_2t_-IsoP, were significantly lower in CRC cases compared to controls in the SWHS and SMHS (*P* = .01) **(Supplementary Table 3)**. The multivariable-adjusted ORs (95% CI) for CRC at the 10th and 90th percentiles of 15-F_2t_-IsoP-M, relative to the median, were 1.11 (1.06-1.17) and 0.83 (0.71-0.97), respectively, with overall *P* < .0001 (**Table 1**). However, this association was not observed in the SCCS **(Table 1)**.

We also compared the ratio of metabolic product 15-F_2t_-IsoP-M to substrate 15-F_2t_-IsoP (RPS-IsoP) between cases and controls. CRC cases tended to have a lower RPS-IsoP compared to controls **(Supplementary Table 3).** Elevated the metabolism ratio was associated with reduced CRC risk (overall *P* = .04, **Table 1**). The multivariable-adjusted ORs (95% CI) for CRC at the 10th and 90th percentiles of RPS-IsoP, relative to the median, were 1.22 (1.04-1.43) and 0.90 (0.82-0.98), respectively.

Unlike the relationship between 5-F_2t_-IsoP and CRC risk weakening over time, the association with the β-oxidation product 15-F_2t_-IsoP-M strengthened with longer follow-up (**Table 2** and **Supplementary** Figures 2 and 3). For example, baseline levels of 15-F_2t_-IsoP-M were not associated with CRC risk diagnosed within 5 years of enrollment (*P* = .14) but were significantly associated with CRC diagnosed between 5 and 9 years (*P* = .04), and even more strongly associated with CRC diagnosed after 9 years (*P* = .01), with *P* for heterogeneity of < .001.

### Results of sensitivity analyses

The robustness of our findings was evaluated through stratified analyses, showing that the inverse and time-dependent association between 5-F_2t_-IsoP and CRC risk remained consistent across sexes and anatomic locations of CRC, although insignificant in some subgroups likely due to the smaller sample sizes (**Supplementary Tables 5-8**). We also evaluated the impact of normalizing inter-batch variations. The results were similar with and without normalization in analyses confined to batches with CV <20% (**Table 1** and **Supplementary Tables 10and 11**) and in analyses involving all study participants (**Supplementary Tables 12and 13**). Additionally, the time-dependent association persisted when finer time stratifications were applied (data not shown).

### Association of CRC risk with composite index of three systemic OxS markers

We formulated a composite index of 8-oxo-dG (DNA), 8-oxo-Guo (RNA) and 5-F_2t_-IsoP (lipid) markers based on the principal component analysis. The inverse association between the index and CRC risk was observed for both the Shanghai and US cohorts **(Supplementary Table 13)** and varied with follow-up time **(Supplementary Table 13 and Supplementary** Figure 4). The index of OxS markers was found to be superior to individual markers alone in predicting CRC risk (**Table 3**). For example, ORs (95% CI) for CRC diagnosed within 5 years of enrollment at the 10th and 90th percentiles of the index, relative to the median, were 2.11 (1.44-3.09) and 0.36 (0.25-0.51), respectively, indicating an approximately 4-fold difference in risk between the two groups, compared to 2-to 3-fold differences in risk between the two groups when using individual markers.

**Table 3.** Multivariable-adjusted odds ratio and confidence intervals for colorectal cancer diagnosed within 5 years of enrollment: comparison of oxidative stress index with individual markers in the SWHS and SMHS.

|  | OR (95% CI) for colorectal cancer risk relative to reference, <sup>a</sup><br>by percentile distribution of biomarkers <sup>b</sup> |  |  |  |  | *P* for<br>overall<br>association | *P* for<br>nonlinear<br>association | Fold difference in<br>OR b/w the 10<sup>th</sup><br>and the 90<sup>th</sup> |
| --- | --- | --- | --- | --- | --- | --- | --- | --- |
|  | 10th | 30th | 50th | 70th | 90th |  |  |  |
| **Index of OxS markers <sup>c</sup>** | 2.11 (1.44, 3.09) | 1.43 (1.20, 1.70) | Reference | 0.62 (0.52, 0.75) | 0.36 (0.25, 0.51) | <.001 | .062 | 3.89 |
| **5-F<sub>2t</sub>-IsoP** | 1.57 (1.26, 1.96) | 1.22 (1.11, 1.33) | Reference | 0.79 (0.71, 0.88) | 0.61 (0.42, 0.89) | <.001 | .018 | 2.20 |
| **8-oxo-dG <sup>d</sup>** | 1.87 (1.39, 2.53) | 1.34 (1.17, 1.54) | Reference | 0.70 (0.61, 0.80) | 0.48 (0.37, 0.63) | <.001 | .02 | 2.95 |
| **8-oxo-Guo <sup>d</sup>** | 1.46 (1.07, 1.98) | 1.19 (1.04, 1.37) | Reference | 0.80 (0.69, 0.93) | 0.57 (0.42, 0.76) | <.001 | .26 | 2.21 |

Abbreviations: 5-F<sub>2t</sub>-IsoP, 5-F<sub>2t</sub>-Isoprostane; 8-oxo-dG, 8-oxo-7,8-dihydro-2'-deoxyguanosine; 8-oxo-Guo, 7,8-dihydro-8-oxo-guanosine; CI, confidence interval; OR, odds ratio; SMHS, Shanghai Men's Health Study; SWHS, Shanghai Women's Health Study.
<sup>a</sup> The OR was estimated using a conditional logistic regression model with restricted cubic spline functions, with the 50th percentile of biomarker in controls as the reference, and adjusted for age at baseline, education, cigarette smoking, alcohol consumption, BMI, physical activity, regular use of vitamin supplements, regular use of aspirin and other nonsteroidal anti-inflammatory drugs, Charlson comorbidity score, family history of colorectal cancer in first-degree relatives, and total energy intake.
<sup>b</sup> Cutoffs were based on the percentile distribution of biomarker levels in controls as detailed in the footnotes of Table 1.
<sup>c</sup> The index was constructed using the principal component analysis for 8-oxo-dG (DNA), 8-oxo-Guo (RNA) and 5-F<sub>2t</sub>-IsoP (lipid) OxS markers.
<sup>d</sup> Details have been reported in MedRxiv. 2025; doi:10.1101/2025.01.21.25320898.

## DISCUSSION

This study presents new supporting evidence for the inverse and time-dependent relationship between systemic OxS and CRC risk. The relationship with systemic lipid peroxidation, along with our recent findings on DNA and RNA OxS markers,^1^ suggests a robust and consistent effect of all three types (DNA, RNA and lipids) of oxidative damages on CRC risk. Individuals at the 10^th^ percentile of these markers, compared to those in the 90^th^ percentile, experienced a 2-to 3-fold increased risk of CRC in the late phase of its development. This association remained robust across stratified analyses by sex and anatomical site of CRC. Importantly, it was consistently observed across three study cohorts, despite differences in baseline characteristics, including racial ethnicity and risk profiles, between the Shanghai and the US cohorts.

A shift in the role of OxS from being pro-tumorigenic to anti-tumorigenic across tumorigenesis has been demonstrated in experimental models.^2–4^ Cancer cells are typically more vulnerable to high OxS than normal cells. Increased OxS, if not controlled, can damage macromolecules vital for cell survival, resulting in senescence or apoptosis, thus acting like a tumor suppressor.^18,19^ Cancer cells under metabolic stress undergo metabolic reprogramming,^20–22^ activating a variety of antioxidant defense mechanisms to mitigate OxS.^4,21,23,24^ Lowering OxS through genetic or pharmacological manipulation in experimental models fosters a survival advantage to cancer cells, promoting further growth and metastasis.^3,25,26^ For example, previous research has demonstrated increased expression of superoxide dismutase, a key antioxidant enzyme, in CRC patients.^27,28^ This increase was observed both in tumor tissues and in circulation and was closely correlated to the severity of CRC.^29,30^

Urinary F_2_-IsoPs, products of free radical-catalyzed peroxidation of arachidonic acid,^31^ have long be characterized as reliable OxS markers.^32–34^ The GC-NICI-MS assay, developed in the 1990s, has been widely used to quantify 15-F_2t_-IsoP and co-eluting isomers. Importantly, 15-F_2t_-IsoP, the most studied F_2_-IsoP, is generated not only via free radical-induced oxidation of arachidonic acid but also via cyclooxygenase-catalyzed oxidation.^17^ In this study, we measured 5-F_2t_-IsoP, an abundant isomer of F_2_-IsoPs that is formed exclusively through free radical-induced oxidation, along with 15-F_2t_-IsoP and its β-oxidation metabolite (15-F_2t_-IsoP-M), allowing us to assess systemic OxS more specifically and accurately and also contextualize the study findings within existing literature.

Prospective investigations on F_2_-IsoPs and CRC risk have been very limited. Two nested case-control studies reported that higher levels of serum hydroperoxides and oxidized low-density lipoprotein were associated with increased CRC risk.^35,36^ However, these two markers are not considered reliable systemic OxS assessment.^10,31^ Additionally, these markers were measured in serum, whereas urine is considered a better matrix for systemic OxS assessment due to low concentrations of material that can artifactually generate or promote lipid peroxidation during sampling and long-term storage.^33,39^ In contrast, a null association between urinary F_2_-IsoPs and colorectal adenoma risk was reported in a cohort study after 10 years of follow-up.^40^ Most previous efforts have employed a cross-sectional design,^41–43^ comparing cancer-free individuals to cancer cases with biological samples collected after diagnosis and often during cancer treatment. This approach is problematic for investigating early molecular events. Additionally, these studies usually involved very small samples (<100 cases),^41–43^ and ELISA method was commonly used, which is not considered a valid substitute for mass spectrometry-based assays.^44^ More importantly, little attention has been paid to the potential time-dependent effect of OxS. To our knowledge, this is the first population-based study focusing on the dynamic relationship between systemic lipid peroxidation and CRC risk over time.

Given that more than two decades can elapse between initiation and clinical emergence of cancer in humans,^45^ individuals diagnosed within the first 5 years of enrollment, as examined in this study, are very likely to have already been in their later phase of cancer development at enrollment. Therefore, the new finding of this study, along with our recent report on DNA and RNA OxS markers,^1^ suggests that the observed systemic OxS reduction likely occurs in the late phase of colorectal tumorigenesis, potentially coinciding with metabolic reprogramming at the primary cancer site. Because we do not fully understand the signaling pathways responsible for the systemic alteration and because this is a new finding, further investigations into the temporal pattern are truly warranted.

We also observed that, unlike unmetabolized 15-F_2t_-IsoP, its β-oxidation metabolite 15-F_2t_-IsoP-M levels or the product-to-substrate ratio (RPS-IsoP) were inversely and time-dependently associated with CRC risk. While the 5-F_2t_-IsoP and CRC association weakened over time, the association with 15-F_2t_-IsoP-M strengthened with extended follow-up. Since levels of its parent compound 15-F_2t_-IsoP were not associated with CRC risk, the relationship with the metabolite 15-F_2t_-IsoP-M may be primarily determined by β-oxidation activity. We recently found that 15-F_2t_-IsoP-M is a better predictor of future weight change than its unmetabolized F_2_-IsoPs.^46^ Additionally, a cohort study reported that elevated 15-F_2t_-IsoP-M levels in urine were associated with reduced type 2 diabetes.^47^ The biological mechanisms underlying this relationship are not fully understood. Alternatively, this could be a chance finding, as it was observed only in the Shanghai cohorts and not replicated in the US cohort.

Our study has several notable strengths, including the two-stage approach involving both the primary and replication analyses, the use of specific biomarkers quantified by mass spectrometric-based methods, and comprehensive analytic approaches to evaluate the association in a time-dependent manner. Unlike previous studies that were often constrained by small sample sizes or retrospective designs, the large sample size and long-term follow-up in the SWHS and SMHS allowed us to investigate alterations in systemic lipid peroxidation with CRC progression, although the number of cases in the SCCS was insufficient for the time-varying effect analysis.

Some limitations or concerns should be mentioned. First, urine samples were collected at a single time point. However, we found that both F_2_-IsoPs and 15-F_2t_-IsoP-M levels remain stable in serial samples collected over a 1-year period,^48^ suggesting that a single measurement reflects an individual’s long-term OxS levels reasonably well. Another concern is the possibility of residual confounding, although a wide range of covariates had already been considered in the study. Additionally, it has been suggested that the systemic OxS and cancer risk association may vary by cancer types.^49–56^ Therefore, further investigations into this time-varying effect for various cancer types are clearly needed.

In conclusion, our study, along with our earlier findings,^1^ suggests that reduced systemic OxS is associated with increased risk of CRC in its late phase of development. This echoes previous RCTs indicating that lowering systemic OxS through antioxidant supplementation could potentially pose risks for high-risk individuals. Therefore, the inverse and time-dependent relationship between OxS and CRC development, if further confirmed, likely has high implications in cancer prevention given the widespread use of antioxidant supplements in this study and many other populations.

## Data Availability

Data described in the article, code book, and analytic codes will be made available upon request pending approval by the data management and sharing committees of the SWHS, SMHS and SCCS.

## Abbreviations

5-F_2t_-IsoP (also known as 5-iPF_2α_-VI), 5-F_2t_-Isoprostane; 15-F_2t_-IsoP (also known as 8-iso-PGF_2a_), 8-oxo-dG, 8-oxo-7,8-dihydro-2’-deoxyguanosine; 8-oxo-Guo, 7,8-dihydro-8-oxo-guanosine; 15-F_2t_-Isoprostane; 15-F_2t_-IsoP-M, 2,3-dinor-5,6-dihydro-15-F_2t_-Isoprostane; CI, confidence interval; CRC, colorectal cancer; GC-NICI-MS, gas chromatography-negative ion chemical ionization-mass spectrometry; F_2_-IsoPs, F_2_-isoprostanes; ng/mg cr, nanograms per milligram of creatinine; OR, odds ratio; OxS, oxidative stress; RCT, randomized controlled trials; RPS-IsoP, the ratio of 15-F_2t_-IsoP-M (metabolic product) to 15-F_2t_-IsoP (substrate); SCCS, Southern Community Cohort Study; SMHS, Shanghai Men’s Health Study; SWHS, Shanghai Women’s Health Study; UPLC-MS/MS, ultra-performance liquid chromatography tandem mass spectrometry.

## Author Contributions

Dr. Yang had full access to all the data in the study and take responsibility for the integrity of the data and the accuracy of the data analysis.

*Concept and design:* Yang, Milne and Chen.

*Acquisition, analysis, or interpretation of data:* All authors.

*Drafting of the manuscript:* Yang.

*Critical revision of the manuscript for important intellectual content:* All authors.

*Statistical analysis:* Yi and Chen.

*Obtained funding:* Yang, Shu, Zheng, and Chen.

*Administrative, technical, or material support:* Yang, Milne, and Chen.

*Supervision:* Yang.

## Additional Contributions

We are grateful to the participants and research staff of the Shanghai Women’s Health Study, the Shanghai Men’s Health Study, and the Southern Community Cohort Study for their contributions to the study. We thank Jie Wu, Regina Courtney and Rodica Cal-Chris for contributions to sample preparation at the Survey and Biospecimen Shared Resources. These individuals were not compensated.

