## Supplemental data for "Lipid peroxidation and colorectal cancer risk: a time-varying relationship"

#### Supplementary Online Content

1. **Supplementary Methods**
2. **Supplementary Table 1.** Baseline characteristics of colorectal cancer cases and matched controls from the SWHS and the SMHS
3. **Supplementary Table 2.** Baseline characteristics of colorectal cancer cases and matched controls from the SCCS
4. **Supplementary Table 3.** Urinary levels of $F_2$-IsoPs in colorectal cancer cases and controls
5. **Supplementary Table 4.** Associations between baseline urinary levels of $F_2$-IsoPs and subsequent risk of colorectal cancer by sex in the SWHS and SMHS
6. **Supplementary Table 5.** Associations between baseline urinary levels of $F_2$-IsoPs and subsequent risk of colorectal cancer by race in the SCCS
7. **Supplementary Table 6.** Associations between baseline urinary levels of $F_2$-IsoPs and subsequent risk of colorectal cancer by time interval between urine sample collection at baseline and cancer diagnosis during follow-up in the SWHS
8. **Supplementary Table 7.** Associations between baseline urinary levels of $F_2$-IsoPs and subsequent risk of colorectal cancer by time interval between urine sample collection at baseline and cancer diagnosis during follow-up in the SMHS

9. **Supplementary Table 8.** Associations between baseline urinary levels of $F_2$-IsoPs and subsequent risk of colorectal cancer in the SWHS and the SMHS, stratified by anatomical sites and by time interval between urine sample collection at baseline and cancer diagnosis during follow-up
10. **Supplementary Table 9.** Associations between baseline urinary levels of $F_2$-IsoPs and subsequent risk of colorectal cancer: analyses confined to batches with CV <20% and without inter-batch normalization
11. **Supplementary Table 10.** Associations between baseline urinary levels of $F_2$-IsoPs and subsequent risk of colorectal cancer stratified by follow-up time: analyses confined to batches with CV <20% and without inter-batch normalization
12. **Supplementary Table 11.** Associations between baseline urinary levels of $F_2$-IsoPs and subsequent risk of colorectal cancer: analyses in all participants without inter-batch normalization
13. **Supplementary Table 12.** Associations between baseline urinary levels of $F_2$-IsoPs and subsequent risk of colorectal cancer stratified by follow-up time: analyses in all participants from the SWHS and SMHS and without inter-batch normalization
14. **Supplementary Table 13.** Associations between the index of DNA, RNA, and lipid OxS markers and subsequent risk of colorectal cancer
15. **Supplementary Figure 1.** Smoothed plot for multivariable-adjusted ORs for colorectal cancer risk according to urinary levels of 15-$F_{2t}$-IsoP by time interval

from baseline to cancer diagnosis

16. **Supplementary Figure 2.** Smoothed plot for multivariable-adjusted ORs for colorectal cancer risk according to urinary levels of 15-$F_{2t}$-IsoP-M by time interval from baseline to cancer diagnosis
17. **Supplementary Figure 3.** Smoothed plot for multivariable-adjusted ORs for colorectal cancer risk according to RPS-IsoP by time interval from baseline to cancer diagnosis
18. **Supplementary Figure 4.** Smoothed plot for multivariable-adjusted ORs for colorectal cancer risk according to OxS marker index in the SWHS and SMHS by time interval from baseline to cancer diagnosis.

### 1. Supplementary Method

#### 1.1. Laboratory Measurements

A stock solution of the isotopically-labeled internal standard ([$^2H_4$]-15-$F_{2t}$-IsoP, Item No. 316350, Cayman Chemical, Ann Arbor, MI USA) was prepared in ethanol. The concentration of the stock solution was determined by comparison with the 15-$F_{2t}$-IsoP MaxSpec© standard (Item No. 25903, Cayman Chemical, Ann Arbor, MI USA).

Urine was thawed on ice. A 0.200mL aliquot was diluted with 0.400mL of a solution of 0.1% formic acid/methanol (95/5, v/v) and 0.040mL 0.1N HCl. The solution was vortexed to mix and to [$^2H_4$]-15-$F_{2t}$-IsoP (1.8ng) was added to each sample. The samples were again mixed and the pH of each was adjusted to pH3 with 0.1N HCl, if required.

The samples were purified by extraction on a Waters HLB 96-well micro-elution plate (Waters Corporation, Milford, MA USA). Sample wells were first washed with methanol (0.200mL x 2) followed by 25% methanol in water (0.200mL x 2). The sample was then loaded on the matrix and washed with 0.400mL of a solution of 0.1% formic acid/methanol (95/5, v/v). The wells were then washed with 0.200mL hexanes. Isoprostanes and metabolites were eluted from the plate with 0.030mL 2-propanol/acetonitrile (50/50, v/v) into a 96-well collection plate containing 0.030mL water in each well.

Samples were analyzed on a Waters Xevo TQ-XS triple quadrupole mass spectrometer connected to a Waters Acquity I-Class UPLC (Waters Corp., Milford,

MA USA). Separation of analytes was obtained using a Waters BEH C18 UPLC column (1.0 x 100mm, 1.8mm) with mobile phase A being 0.01% formic acid in water and mobile phase B acetonitrile. Isoprostanes and metabolites were separated using a gradient elution beginning with 20% B going to 95% B at a flow rate of 0.100mL/min (see Gradient Table). The mass spectrometer was operated using multiple reaction monitoring (MRM) in the negative ion mode with argon as the collision gas. Instrument control and data acquisition utilized MassLynx V4.2; integration and quantitation used TargetLynx 4.2. The limit of detection is 0.050 ng/mL; CV, <8%; accuracy, 94%.

**UPLC Conditions**

| | |
|---|---|
| **Column Temp** | 35$^{o}$C |
| **Sample Temp** | 8$^{o}$C |
| **Inj Mode** | Partial Loop |
| **Inj Loop** | 10μL |
| **Inj Vol** | 5μL |

**UPLC Gradient**

| Time (min) | Flow Rate (mL/min) | %A | %B | Curve |
|---|---|---|---|---|
| Initial | 0.100 | 80 | 20 | Initial |
| 0.10 | 0.100 | 80 | 20 | 6 |
| 5.00 | 0.100 | 60 | 40 | 6 |
| 6.00 | 0.100 | 5 | 95 | 6 |
| 8.50 | 0.100 | 5 | 95 | 6 |
| 8.60 | 0.100 | 70 | 30 | 6 |
| 11.00 | 0.100 | 70 | 30 | 6 |

**Mass Spectrometer Settings**

| | |
|---|---|
| **Operation Mode** | ESI negative |
| **Capillary** | 1 kV |
| **Desolvation Temp** | 500 V |
| **Desolvation Gas** | 1000 L/hr |
| **Cone Gas** | 150 L/hr |
| **Nebuliser** | 7 bar |

**Mass-to-charge ratios for $F_2$-IsoPs**

| Analyte | Internal Std | Precursor Ion (m/z) | Product Ion (m/z) | Cone (Volts) | Collision Energy | Dwell time (seconds) |
|---|---|---|---|---|---|---|
| 2,3-dinor-5,6-dihydro-15-$F_{2t}$-IsoP | 15-$F_{2t}$-IsoP-$d_4$ | 327.1 | 283.1 | 40 | 24 | 0.022 |
| 15-series-$F_2$-IsoPs | 15-$F_{2t}$-IsoP-$d_4$ | 353.1 | 193.1 | 40 | 24 | 0.022 |
| 5-series-$F_2$-IsoPs | 15-$F_{2t}$-IsoP-$d_4$ | 353.102 | 115.1 | 40 | 24 | 0.022 |

#### 1.2. Correction of batch effects and sensitivity analysis

Both intra-batch and inter-batch variabilities may contribute to the uncertainty of the research results. In this study, the test samples were delivered to the laboratory subsequently by 5 sampling groups, containing several hundreds to over a thousand study samples in each. Generally, about 30 samples including 4 quality control (QC) samples were measured in each intra-day batch. The entire measurement was completed over a span of approximately 2 years, with a minor refinement in the laboratory method. We carefully evaluated the potential influence of these issues and incorporated intra-batch and inter-batch variations, QC assessment, and methodology refinement into our downstream analyses.

The intra-batch effects were investigated by calculating the coefficient of variation (CV) of QC samples within each batch (n=4, accounting for ~13% of study samples measured in each intra-day batch). The CVs for selected analytes in our laboratory are typically <8%. Outliers with high CV (>20%) were excluded from the main analysis. Sensitivity analyses involving all participants were also performed.

To minimize the potential impact of inter-batch variation, samples from each case-control pair were analyzed in the same assay batch with a random and blind arrangement. Conditional logistic regression models were used, stratified by matched case-control sets, which greatly minimized the potential influence of inter-batch variations on the results.

We applied the correction of inter-batch effects by normalization. We first calculated a QC sample-based batch-specific ratio (QBR) as a ratio between the QC sample median within each batch and the median of QC samples from all batches. Next, we normalized the biomarker measurement by dividing the corresponding batch-specific ratio, which will be used in the downstream analyses. This normalization method assumes the QBR represents the inter-batch effect, and the same QC sample was used in all batches. However, two QC samples were used in this study. To further normalize the multiple QC sample effects, another ratio factor was derived as the ratio between the medians of two QC samples (QR) and multiplied by QBR.

This two-step transformation leads to the following normalization algorithm. Let

$$Normalized_{ik} = Sample_{ik} \times QBR_{ik} \times QR_k,$$

$$QBR_{ik} = \frac{X_{.k}}{X_{ik}},\ i = 1 \ldots n_k,\ k = 1,2$$

where $Sample_{ik}$ stands for original biomarker measurement, $i$ stands for batch, $X_{ik}$ is the median of QC sample in $i^{th}$ batch using $k^{th}$ QC sample, and $X_{.k}$ is the median from all batches using $k^{th}$ QC sample. In this study, $QR_k$ equals to ratio between the median of 1st QC samples and the median of $k^{th}$ QC samples, and $QR_1$ equals to 1. Normalized biomarker measurement $Normalized_{ik}$ was then used in the downstream analysis.

Model analyses without correcting intra- and inter-batch effects (excluding batches with CV>20% and normalizing assay results using QC median) were also performed.

**Supplementary Table 1. Baseline characteristics of colorectal cancer cases and matched controls from the SWHS and the SMHS**

| Characteristics | All | | | Females in SWHS | | | Males in SMHS | | |
|---|---|---|---|---|---|---|---|---|---|
| | Cases (n = 1938) | Controls (n = 1938) | *P* value [a] | Cases (n = 1035) | Controls (n = 1035) | *P* value [a] | Cases (n = 903) | Controls (n = 903) | *P* value [a] |
| Age (year), mean (SD) | 59.4 (9.2) | 59.2 (9.2) | 0.741 | 57.5 (8.8) | 57.5 (8.9) | 0.981 | 61.5 (9.2) | 61.3 (9.2) | 0.630 |
| Education, high school and above, n (%) | 865 (44.6) | 844 (43.6) | 0.523 | 361 (34.9) | 342 (33.0) | 0.402 | 504 (55.8) | 502 (55.6) | 0.969 |
| Alcohol consumption, n (%) | 334 (17.2) | 346 (17.9) | 0.644 | 22 (2.1) | 26 (2.5) | 0.661 | 312 (34.6) | 320 (35.4) | 0.734 |
| Cigarette smoking, n (%) | 605 (31.2) | 628 (32.4) | 0.453 | 29 (2.8) | 38 (3.7) | 0.327 | 576 (63.8) | 590 (65.3) | 0.526 |
| Family history of colorectal cancer, n (%) | 65 (3.4) | 36 (1.9) | 0.0052 | 38 (3.7) | 19 (1.8) | 0.023 | 27 (3.0) | 17 (1.9) | 0.178 |
| Postmenopausal, n (%) | NA | NA | NA | 749 (72.4) | 759 (73.3) | 0.687 | NA | NA | NA |
| Physical activity (MET-h/wk), mean (SD) | 89.0 (44.0) | 88.8 (47.3) | 0.275 | 108.2 (41.5) | 109.7 (45.8) | 0.913 | 67.1 (35.7) | 64.7 (36.3) | 0.098 |
| Body mass index (kg/m$^2$), mean (SD) | 24.4 (3.3) | 24.2 (3.4) | 0.069 | 24.5 (3.4) | 24.6 (3.6) | 0.549 | 24.3 (3.2) | 23.7 (3.0) | <0.001 |
| Use of vitamin supplements, n (%) | 348 (18.0) | 318 (16.4) | 0.228 | 189 (18.3) | 185 (17.9) | 0.864 | 159 (17.6) | 133 (14.7) | 0.112 |
| Use of aspirin and other NSAIDs, n (%) | 134 (6.9) | 115 (5.9) | 0.241 | 20 (1.9) | 31 (3.0) | 0.165 | 114 (12.6) | 84 (9.3) | 0.031 |
| Total energy intake (kcal/d), mean (SD) | 1775.6 (439.8) | 1786.4 (475.0) | 0.950 | 1673.5 (394.1) | 1693.9 (428.1) | 0.450 | 1892.6 (460.1) | 1892.5 (503.4) | 0.557 |
| Charlson comorbidity index, mean (SD) | 0.4 (0.9) | 0.4 (0.8) | 0.967 | 0.3 (0.7) | 0.3 (0.8) | 0.061 | 0.6 (1.0) | 0.5 (0.9) | 0.065 |

Abbreviations: SWHS, Shanghai Women's Health Study; SMHS, Shanghai Men's Health Study; case, study participant who had developed colorectal cancer during follow-up; control, study participant who remained cancer free; MET, metabolic equivalent; NSAID, nonsteroidal anti-inflammatory drug; NA, not applicable.

[a] Wilcoxon rank sum test was used for continuous variables and chi-square test for categorical variables.

**Supplementary Table 2. Baseline characteristics of colorectal cancer cases and matched controls from the SCCS**

| Characteristics | All | | | African Americans | | | European Americans | | |
|---|---|---|---|---|---|---|---|---|---|
| | Cases (n = 285) | Controls (n = 570) | *P* value [a] | Cases (n = 192) | Controls (n = 384) | *P* value [a] | Cases (n = 93) | Controls (n = 186) | *P* value [a] |
| Age (year), mean (SD) | 55.1 (8.8) | 55.0 (8.7) | 0.846 | 54.6 (8.8) | 54.4 (8.7) | 0.793 | 56.1 (8.7) | 56.1 (8.6) | 0.982 |
| Education, high school and above, n (%) | 181 (63.5) | 359 (63.0) | 0.895 | 115 (59.9) | 231 (60.2) | 0.995 | 66 (71.0) | 128 (68.8) | 0.821 |
| Alcohol consumption, n (%) | 237 (83.2) | 478 (83.9) | 0.997 | 165 (85.9) | 325 (84.6) | 0.827 | 72 (77.4) | 153 (82.3) | 0.646 |
| Cigarette smoking, n (%) | 180 (63.2) | 371 (65.1) | 0.736 | 116 (60.4) | 243 (63.3) | 0.629 | 64 (68.8) | 128 (68.8) | 0.998 |
| Family history of colorectal cancer, n (%) | 34 (11.9) | 43 (7.5) | 0.044 | 18 (9.4) | 26 (6.8) | 0.345 | 16 (17.2) | 17 (9.1) | 0.074 |
| Gender, male (%) | 112 (39.3) | 224 (39.3) | 0.993 | 80 (41.7) | 160 (41.7) | 0.994 | 32 (34.4) | 64 (34.4) | 0.992 |
| Physical activity (MET-h/wk), mean (SD) | 160.0 (136.5) | 160.7 (140.5) | 0.909 | 156.5 (142.4) | 152.9 (141.8) | 0.715 | 167.4 (123.6) | 176.8 (136.9) | 0.705 |
| Body mass index ($kg/m^2$), mean (SD) | 30.7 (7.9) | 31.2 (7.6) | 0.234 | 30.1 (7.4) | 31.2 (8.0) | 0.142 | 31.8 (8.7) | 31.1 (6.9) | 0.873 |
| Use of vitamin supplements, n (%) | 116 (40.7) | 249 (43.7) | 0.491 | 78 (40.6) | 156 (40.6) | 0.990 | 38 (40.9) | 93 (50.0) | 0.258 |
| Use of aspirin and other NSAIDs, n (%) | 32 (11.2) | 73 (12.8) | 0.590 | 21 (10.9) | 50 (13.0) | 0.567 | 11 (11.8) | 23 (12.4) | 0.995 |
| Total energy intake (kcal/d), mean (SD) | 2596.9 (1540.5) | 2547.2 (1439.6) | 0.892 | 2726.8 (1591.5) | 2692.5 (1524.6) | 0.871 | 2334.3 (1403.7) | 2262.3 (1210.5) | 0.863 |
| Charlson comorbidity index, mean (SD) | 2.0 (1.5) | 2.1 (1.4) | 0.309 | 1.9 (1.5) | 2.1 (1.4) | 0.108 | 2.3 (1.6) | 2.2 (1.4) | 0.540 |

Abbreviations: SCCS, Southern Community Cohort Study; case, study participant who had developed colorectal cancer; control, study participant who remained cancer free; MET, metabolic equivalent; NSAID, nonsteroidal anti-inflammatory drug.

[a] Wilcoxon rank sum test was used for continuous variables and chi-square test for categorical variables

**Supplementary Table 3. Urinary levels (ng/mg Cr) of $F_2$-IsoPs in colorectal cancer cases and controls from the SWHS, SMHS and SCCS**

| | Cases | | Controls | | Difference b/w case and control [a] | *P* value [b] |
|---|---|---|---|---|---|---|
| | Geometric mean | 95% CI | Geometric mean | 95% CI | | |
| **SWHS and SMHS** | | | | | | |
| 5-$F_{2t}$-IsoP | 6.22 | (6.02, 6.43) | 6.76 | (6.54, 6.99) | -8.0% | < 0.001 |
| 15-F2t-IsoP | 2.13 | (2.06, 2.21) | 2.16 | (2.09, 2.24) | -1.4% | 0.602 |
| 15-$F_{2t}$-IsoP-M | 9.07 | (8.73, 9.42) | 9.73 | (9.37, 10.10) | -6.8% | 0.007 |
| RPS-IsoP | 556% | (534%, 578%) | 588% | (565%, 612%) | -5.4% | 0.043 |
| **SCCS** | | | | | | |
| 5-$F_{2t}$-IsoP | 7.27 | (6.58, 8.03) | 7.93 | (7.39, 8.52) | -8.3% | 0.165 |
| 15-F2t-IsoP | 1.67 | (1.54, 1.82) | 1.75 | (1.65, 1.86) | -4.6% | 0.361 |
| 15-$F_{2t}$-IsoP-M | 14.09 | (12.83, 15.49) | 14.76 | (13.80, 15.78) | -4.5% | 0.440 |
| RPS-IsoP | 1320% | (1200%, 1453%) | 1318% | (1231%, 1410%) | 0.2% | 0.974 |

Abbreviations: 5-F2t-IsoP, 5-F2t-Isoprostane; 15-F2t-IsoP, 15-F2t-Isoprostane; 15-F2t-IsoP-M, 2,3-Dinor-5,6-dihydro-15-F2t-Isoprostane; RPS-IsoP, ratio of 15-$F_{2t}$-IsoP-M (β-oxidation product) to 15-$F_{2t}$-IsoP (substrate); SWHS, Shanghai Women's Health Study; SMHS, Shanghai Men's Health Study; SCCS, Southern Community Cohort Study; All biomarker values are centered by batch median.

Total sizes of case: control in the SWHS and SMHS: 1938:1938; in the SCCS: 285: 570.

[a] Difference (%) = (geometric mean of biomarker in case – geometric mean of biomarker in control)/geometric mean of biomarker in control.

[b] Derived from paired *t*-tests for SWHS and SMHS and from mixed-effects models for SCCS due to 1:2 matching.

**Supplementary Table 4. Associations between baseline urinary levels of $F_2$-IsoPs and subsequent risk of colorectal cancer by sex (cohort) in the SWHS and SMHS**

| | Number of pairs [c] | OR (95% CI) for colorectal cancer risk relative to reference, [a] by percentile distribution of biomarkers [b] | | | | | *P* for overall association | *P* for nonlinear association |
|---|---|---|---|---|---|---|---|---|
| | | 10th | 30th | 50th | 70th | 90th | | |
| **In the SWHS (Females)** | | | | | | | | |
| 5-$F_{2t}$-IsoP | 1013 | 1.07 (0.99, 1.17) | 1.04 (0.99, 1.10) | Reference | 0.92 (0.84, 1.02) | 0.78 (0.57, 1.06) | 0.262 | 0.518 |
| 15-F2t-IsoP | 1013 | 0.98 (0.93, 1.03) | 1.00 (0.99, 1.02) | Reference | 0.96 (0.91, 1.01) | 0.85 (0.71, 1.02) | 0.100 | 0.050 |
| 15-$F_{2t}$-IsoP-M | 1035 | 1.11 (0.99, 1.24) | 1.06 (0.99, 1.13) | Reference | 0.92 (0.83, 1.02) | 0.80 (0.60, 1.06) | 0.031 | 0.657 |
| RPS-IsoP | 1013 | 1.19 (0.96, 1.48) | 1.08 (0.99, 1.18) | Reference | 0.93 (0.86, 0.99) | 0.83 (0.72, 0.95) | 0.027 | 0.398 |
| **In the SMHS (Males)** | | | | | | | | |
| 5-F2t-IsoP | 765 | 1.27 (1.11, 1.46) | 1.12 (1.05, 1.19) | Reference | 0.88 (0.82, 0.94) | 0.80 (0.70, 0.92) | 0.001 | 0.009 |
| 15-F2t-IsoP | 604 | 1.06 (0.97, 1.17) | 1.02 (0.99, 1.05) | Reference | 0.96 (0.90, 1.02) | 0.95 (0.86, 1.07) | 0.409 | 0.196 |
| 15-F2t-IsoP-M | 884 | 1.15 (1.05, 1.26) | 1.07 (1.02, 1.11) | Reference | 0.94 (0.89, 1.00) | 0.86 (0.71, 1.05) | 0.005 | 0.179 |
| RPS-IsoP | 586 | 1.14 (0.93, 1.40) | 1.05 (0.97, 1.14) | Reference | 0.96 (0.90, 1.03) | 0.94 (0.85, 1.04) | 0.302 | 0.203 |

Abbreviations: 5-F2t-IsoP, 5-F2t-Isoprostane; 15-F2t-IsoP, 15-F2t-Isoprostane; 15-F2t-IsoP-M, 2,3-Dinor-5,6-dihydro-15-F2t-Isoprostane; RPS-IsoP, ratio of 15-F2t-IsoP-M (metabolic product) to 15-F2t-IsoP (substrate); CI, confidence interval; ng/mg Cr, nanograms per milligram of creatinine; OR, odds ratio; SMHS, Shanghai Men's Health Study; SWHS, Shanghai Women's Health Study.

[a] Multivariable ORs estimated using a conditional logistic regression model with restricted cubic-spline functions, using the 50th percentile of biomarkers in controls as the reference; and adjusted for age at baseline, education, cigarette smoking, alcohol consumption, BMI, physical activity, menopausal status (for female participants), regular use of vitamin supplements, regular use of aspirin and other nonsteroidal anti-inflammatory drugs, Charlson comorbidity score, family history of colorectal cancer in first-degree relatives, and total energy intake.

[b] Cutoffs were based on the percentile distribution of levels in controls— for 5-$F_{2t}$-IsoP (ng/mg Cr): 1.00 (10th), 1.56 (30th), 2.02 (50th), 2.69 (70th), and 4.28 (90th) in the SWHS; and 1.81 (10th), 2.97 (30th), 4.41 (50th), 6.23 (70th) and 9.64 (90th) in the SMHS; for 15-$F_{2t}$-IsoP (ng/mg Cr): 0.59 (10th), 0.91 (30th), 1.00 (50th), 1.34 (70th), and 2.26 (90th) in the SWHS, and 0.86 (10th), 1.07 (30th), 1.75 (50th), 2.65 (70th) and 4.58 (90th) in the SMHS. for 15-$F_{2t}$-IsoP-M (ng/mg Cr): 4.46 (10th), 6.53 (30th), 8.83 (50th), 12.83 (70th), and 21.88 (90th) in the SWHS; and 4.41 (10th), 9.12 (30th), 13.21 (50th), 18.48 (70th) and 28.79 (90th) in the SMHS; for RPS-IsoP: 422.7% (10th), 649.7% (30th), 853.0% (50th), 1113.5% (70th), and 1770.1% (90th) in the SWHS, and 233.1% (10th), 465.6% (30th), 770.1% (50th), 1180.8% (70th) and 1955.3% (90th) in the SMHS.

[c] The number of case-control pairs in the analysis after exclusion of batches with CV >20%, with the case-control ratio of 1:1.

**Supplementary Table 5. Associations between baseline urinary levels of $F_2$-IsoPs and subsequent risk of colorectal cancer by race in the SCCS**

| | Number of pairs [c] | OR (95% CI) for colorectal cancer risk relative to reference, [a] by percentile distribution of biomarkers [b] | | | | | *P* for overall association | *P* for nonlinear association |
|---|---|---|---|---|---|---|---|---|
| | | 10th | 30th | 50th | 70th | 90th | | |
| **African Americans** | | | | | | | | |
| 5-$F_{2t}$-IsoP | 192 | 1.17 (0.93, 1.47) | 1.09 (0.96, 1.23) | Reference | 0.95 (0.88, 1.03) | 0.97 (0.76, 1.24) | 0.397 | 0.212 |
| 15-$F_{2t}$-IsoP | 192 | 1.05 (0.94, 1.18) | 1.01 (0.99, 1.04) | Reference | 0.98 (0.94, 1.01) | 0.92 (0.80, 1.07) | 0.466 | 0.657 |
| 15-$F_{2t}$-IsoP-M | 192 | 1.13 (1.00, 1.28) | 1.03 (0.98, 1.08) | Reference | 1.01 (0.94, 1.09) | 1.05 (0.82, 1.35) | 0.064 | 0.029 |
| RPS-IsoP | 192 | 1.15 (0.76, 1.75) | 1.07 (0.91, 1.26) | Reference | 0.92 (0.82, 1.03) | 0.78 (0.57, 1.07) | 0.275 | 0.923 |
| **European Americans** | | | | | | | | |
| 5-$F_{2t}$-IsoP | 92 | 1.33 (0.99, 1.78) | 1.18 (0.99, 1.40) | Reference | 0.86 (0.72, 1.03) | 0.91 (0.66, 1.25) | 0.075 | 0.030 |
| 15-$F_{2t}$-IsoP | 92 | 1.03 (0.81, 1.32) | 1.01 (0.94, 1.08) | Reference | 0.99 (0.89, 1.10) | 1.01 (0.80, 1.28) | 0.924 | 0.740 |
| 15-$F_{2t}$-IsoP-M | 92 | 0.91 (0.68, 1.21) | 0.96 (0.86, 1.07) | Reference | 1.04 (0.93, 1.17) | 1.11 (0.75, 1.64) | 0.739 | 0.754 |
| RPS-IsoP | 92 | 0.74 (0.39, 1.43) | 0.88 (0.69, 1.14) | Reference | 1.11 (0.94, 1.30) | 1.23 (0.83, 1.83) | 0.461 | 0.630 |

Abbreviations: 5-$F_{2t}$-IsoP, 5-$F_{2t}$-Isoprostane; 15-$F_{2t}$-IsoP, 15-$F_{2t}$-Isoprostane; 15-$F_{2t}$-IsoP-M, 2,3-Dinor-5,6-dihydro-15-$F_{2t}$-Isoprostane; RPS-IsoP, ratio of 15-F2t-IsoP-M (metabolic product) to 15-F2t-IsoP (substrate); CI, confidence interval; ng/mg Cr, nanograms per milligram of creatinine; OR, odds ratio; SCCS, Southern Community Cohort Study.

[a] Multivariable ORs estimated using a conditional logistic regression model with restricted cubic-spline functions, using the 50th percentile of biomarkers in controls as the reference; and adjusted for age at baseline, education, cigarette smoking, alcohol consumption, BMI, physical activity, menopausal status (for female participants), regular use of vitamin supplements, regular use of aspirin and other nonsteroidal anti-inflammatory drugs, Charlson comorbidity score, family history of colorectal cancer in first-degree relatives, and total energy intake.

[b] Cutoffs were based on the percentile distribution of biomarker levels in controls— for 5-$F_{2t}$-IsoP (ng/mg Cr): 0.69 (10th), 1.07 (30th), 1.70 (50th), 2.73 (70th), and 5.05 (90th) in African Americans in SCCS; and 0.94 (10th), 1.61 (30th), 2.42 (50th), 4.04 (70th) and 6.78 (90th) in European Americans in SCCS; for 15-$F_{2t}$-IsoP (ng/mg Cr): 0.59 (10th), 0.91 (30th), 1.00 (50th), 1.34 (70th), and 2.26 (90th) in African Americans in SCCS, and 0.57 (10th), 1.00 (30th), 1.29 (50th), 1.78 (70th) and 3.08 (90th) in European Americans in SCCS. for 15-$F_{2t}$-IsoP-M (ng/mg Cr): 6.33 (10th), 10.48 (30th), 13.94 (50th), 19.09 (70th), and 29.15 (90th) in African Americans in SCCS; and 7.06 (10th), 11.34 (30th), 16.75 (50th), 24.17 (70th) and 35.27 (90th) in European Americans in SCCS; for RPS-IsoP: 517.8% (10th), 982.6% (30th), 1412.6% (50th), 1796.7% (70th), and 2625.8% (90th) in African Americans in SCCS, and 472.5% (10th), 922.3% (30th), 1252.5% (50th), 1789.6% (70th) and 2522.7% (90th) in European Americans in SCCS.

[c] The number of case-control pairs with the case-control ratio of 1:2. All intra-batch CVs were <20%.

**Supplementary Table 6. Associations between baseline urinary levels of $F_2$-IsoPs and subsequent risk of colorectal cancer by time interval between urine sample collection at baseline and cancer diagnosis during follow-up in the SWHS [a]**

| | Number of pairs [c] | OR (95% CI) for colorectal cancer risk relative to reference, [a] by percentile distribution of biomarkers [b] | | | | | *P* for overall association | *P* for nonlinear association |
|---|---|---|---|---|---|---|---|---|
| | | 10th | 30th | 50th | 70th | 90th | | |
| **Cases diagnosed <5 years after enrollment** | | | | | | | | |
| 5-$F_{2t}$-IsoP | 184 | 1.27 (1.03, 1.56) | 1.13 (1.01, 1.26) | Reference | 0.83 (0.71, 0.99) | 0.52 (0.29, 0.95) | 0.081 | 0.435 |
| 15-$F_{2t}$-IsoP | 184 | 1.01 (0.92, 1.12) | 1.00 (0.98, 1.02) | Reference | 1.01 (0.93, 1.09) | 1.04 (0.76, 1.42) | 0.886 | 0.622 |
| 15-$F_{2t}$-IsoP-M | 185 | 1.45 (0.99, 2.12) | 1.22 (0.99, 1.50) | Reference | 0.71 (0.51, 1.01) | 0.32 (0.10, 1.03) | 0.135 | 0.369 |
| RPS-IsoP | 184 | 0.87 (0.54, 1.41) | 0.97 (0.81, 1.16) | Reference | 0.95 (0.83, 1.08) | 0.63 (0.35, 1.14) | 0.275 | 0.244 |
| **Cases diagnosed 5-9 years after enrollment** | | | | | | | | |
| 5-$F_{2t}$-IsoP | 214 | 1.12 (0.90, 1.39) | 1.09 (0.97, 1.22) | Reference | 0.86 (0.71, 1.04) | 0.58 (0.30, 1.16) | 0.115 | 0.038 |
| 15-$F_{2t}$-IsoP | 214 | 0.95 (0.87, 1.03) | 1.00 (0.99, 1.01) | Reference | 0.98 (0.94, 1.03) | 0.91 (0.76, 1.10) | 0.197 | 0.075 |
| 15-$F_{2t}$-IsoP-M | 216 | 1.04 (0.91, 1.19) | 1.02 (0.95, 1.10) | Reference | 0.97 (0.86, 1.09) | 0.89 (0.59, 1.33) | 0.376 | 0.391 |
| RPS-IsoP | 214 | 1.55 (1.04, 2.29) | 1.18 (1.01, 1.37) | Reference | 0.94 (0.85, 1.04) | 1.16 (0.75, 1.80) | 0.081 | 0.027 |
| **Cases diagnosed >9 years after enrollment** | | | | | | | | |
| 5-$F_{2t}$-IsoP | 615 | 1.02 (0.96, 1.08) | 1.00 (0.98, 1.03) | Reference | 1.00 (0.95, 1.05) | 1.00 (0.84, 1.18) | 0.746 | 0.543 |
| 15-$F_{2t}$-IsoP | 615 | 0.96 (0.89, 1.03) | 1.00 (0.99, 1.01) | Reference | 1.00 (0.97, 1.03) | 0.94 (0.83, 1.06) | 0.248 | 0.110 |
| 15-$F_{2t}$-IsoP-M | 614 | 1.06 (0.97, 1.17) | 1.03 (0.98, 1.09) | Reference | 0.95 (0.86, 1.05) | 0.84 (0.59, 1.20) | 0.204 | 0.764 |
| RPS-IsoP | 614 | 1.14 (0.93, 1.41) | 1.06 (0.97, 1.17) | Reference | 0.94 (0.87, 1.01) | 0.83 (0.70, 0.98) | 0.089 | 0.545 |

Abbreviations: 5-$F_{2t}$-IsoP, 5-$F_{2t}$-Isoprostane; 15-$F_{2t}$-IsoP, 15-$F_{2t}$-Isoprostane; 15-$F_{2t}$-IsoP-M, 2,3-Dinor-5,6-dihydro-15-$F_{2t}$-Isoprostane; RPS-IsoP, ratio of 15-F2t-IsoP-M (metabolic product) to 15-F2t-IsoP (substrate); CI, confidence interval; ng/mg Cr, nanograms per milligram of creatinine; OR, odds ratio; SWHS, Shanghai Women's Health Study.

[a] Multivariable ORs estimated using a conditional logistic regression model with restricted cubic-spline functions, stratified by the time interval between the baseline sample collection and cancer diagnosis during follow-up, using the 50th percentile of biomarkers in controls as the reference; and multivariable adjusted for age at baseline, education, cigarette smoking, alcohol consumption, BMI, physical activity, menopausal status, regular use of vitamin supplements, regular use of aspirin and other nonsteroidal anti-inflammatory drugs, Charlson comorbidity score, family history of colorectal cancer in first-degree relatives, and total energy intake.

[b] Cutoffs were based on the percentile distribution of levels in controls— for 5-$F_{2t}$-IsoP (ng/mg Cr): 1.00 (10th), 1.56 (30th), 2.02 (50th), 2.69 (70th), and 4.28 (90th) in the SWHS. for 15-$F_{2t}$-IsoP (ng/mg Cr): 0.59 (10th), 0.91 (30th), 1.00 (50th), 1.34 (70th), and 2.26 (90th) in the SWHS. for 15-$F_{2t}$-IsoP-M (ng/mg Cr): 4.46 (10th), 6.53 (30th), 8.83 (50th), 12.83 (70th), and 21.88 (90th) in the SWHS. for RPS-IsoP: 422.7% (10th), 649.7% (30th), 853.0% (50th), 1113.5% (70th), and 1770.1% (90th) in the SWHS.

[c] The number of case-control pairs after exclusion of batches with CV >20%.

**Supplementary Table 7 Associations between baseline urinary levels of $F_2$-IsoPs and subsequent risk of colorectal cancer by time interval between urine sample collection at baseline and cancer diagnosis during follow-up in the SMHS**

| | Number of pairs [c] | OR (95% CI) for colorectal cancer risk relative to reference, [a] by percentile distribution of biomarkers [b] | | | | | *P* for overall association | *P* for nonlinear association |
|---|---|---|---|---|---|---|---|---|
| | | 10th | 30th | 50th | 70th | 90th | | |
| **Cases diagnosed <5 years after enrollment** | | | | | | | | |
| 5-$F_{2t}$-IsoP | 203 | 2.14 (1.44, 3.18) | 1.38 (1.18, 1.61) | Reference | 0.83 (0.72, 0.96) | 0.73 (0.43, 1.24) | < 0.001 | 0.017 |
| 15-$F_{2t}$-IsoP | 183 | 1.12 (0.90, 1.39) | 1.08 (0.93, 1.26) | Reference | 0.96 (0.87, 1.05) | 0.95 (0.77, 1.16) | 0.601 | 0.364 |
| 15-$F_{2t}$-IsoP-M | 226 | 1.10 (0.88, 1.37) | 1.01 (0.92, 1.11) | Reference | 1.02 (0.88, 1.19) | 1.07 (0.69, 1.67) | 0.490 | 0.283 |
| RPS-IsoP | 165 | 1.10 (0.73, 1.66) | 1.05 (0.85, 1.29) | Reference | 0.95 (0.74, 1.20) | 0.91 (0.59, 1.40) | 0.635 | 0.628 |
| **Cases diagnosed 5-9 years after enrollment** | | | | | | | | |
| 5-$F_{2t}$-IsoP | 255 | 1.52 (1.15, 2.02) | 1.19 (1.06, 1.34) | Reference | 0.91 (0.85, 0.98) | 0.91 (0.75, 1.11) | 0.011 | 0.011 |
| 15-$F_{2t}$-IsoP | 144 | 1.40 (1.07, 1.83) | 1.26 (1.05, 1.53) | Reference | 0.90 (0.81, 1.00) | 0.96 (0.73, 1.26) | 0.053 | 0.021 |
| 15-$F_{2t}$-IsoP-M | 323 | 1.18 (1.00, 1.41) | 1.07 (1.00, 1.14) | Reference | 0.93 (0.84, 1.03) | 0.81 (0.60, 1.10) | 0.101 | 0.711 |
| RPS-IsoP | 144 | 0.88 (0.54, 1.44) | 0.94 (0.73, 1.20) | Reference | 1.08 (0.80, 1.47) | 1.15 (0.68, 1.95) | 0.832 | 0.619 |
| **Cases diagnosed >9 years after enrollment** | | | | | | | | |
| 5-$F_{2t}$-IsoP | 307 | 1.09 (0.89, 1.33) | 1.08 (0.97, 1.20) | Reference | 0.87 (0.73, 1.04) | 0.64 (0.37, 1.12) | 0.291 | 0.309 |
| 15-$F_{2t}$-IsoP | 277 | 0.96 (0.84, 1.11) | 0.98 (0.89, 1.07) | Reference | 0.99 (0.90, 1.10) | 0.98 (0.72, 1.34) | 0.851 | 0.581 |
| 15-$F_{2t}$-IsoP-M | 335 | 1.35 (1.06, 1.71) | 1.12 (1.02, 1.23) | Reference | 0.89 (0.76, 1.03) | 0.72 (0.47, 1.12) | 0.035 | 0.534 |
| RPS-IsoP | 277 | 1.45 (0.95, 2.23) | 1.22 (0.97, 1.53) | Reference | 0.75 (0.53, 1.04) | 0.64 (0.37, 1.11) | 0.166 | 0.070 |

Abbreviations: 5-F2t-IsoP, 5-F2t-Isoprostane; 15-F2t-IsoP, 15-F2t-Isoprostane; 15-F2t-IsoP-M, 2,3-Dinor-5,6-dihydro-15-F2t-Isoprostane; RPS-IsoP, ratio of 15-F2t-IsoP-M (metabolic product) to 15-F2t-IsoP (substrate); CI, confidence interval; ng/mg Cr, nanograms per milligram of creatinine; OR, odds ratio; SMHS, Shanghai Men's Health Study.

[a] Multivariable ORs estimated using a conditional logistic regression model with restricted cubic-spline functions, stratified by the time interval between the baseline sample collection and cancer diagnosis during follow-up, using the 50th percentile of biomarkers in controls as the reference; and multivariable adjusted for age at baseline, education, cigarette smoking, alcohol consumption, BMI, physical activity, regular use of vitamin supplements, regular use of aspirin and other nonsteroidal anti-inflammatory drugs, Charlson comorbidity score, family history of colorectal cancer in first-degree relatives, and total energy intake.

[b] Cutoffs were based on the percentile distribution of levels in controls— for 5-$F_{2t}$-IsoP (ng/mg Cr): 1.81 (10th), 2.97 (30th), 4.41 (50th), 6.23 (70th) and 9.64 (90th) in the SMHS; for 15-$F_{2t}$-IsoP (ng/mg Cr): 0.86 (10th), 1.07 (30th), 1.75 (50th), 2.65 (70th) and 4.58 (90th) in the SMHS. for 15-$F_{2t}$-IsoP-M (ng/mg Cr): 4.41 (10th), 9.12 (30th), 13.21 (50th), 18.48 (70th) and 28.79 (90th) in the SMHS; for RPS-IsoP: 233.1% (10th), 465.6% (30th), 770.1% (50th), 1180.8% (70th) and 1955.3% (90th) in the SMHS.

[c] The number of case-control pairs after exclusion of batches with CV > 20%.

**Supplementary Table 8. Associations between baseline urinary levels of $F_2$-IsoPs and subsequent risk of colorectal cancer in the SWHS and the SMHS, stratified by anatomical sites and by time interval between urine sample collection at baseline and cancer diagnosis during follow-up**

| | Number of pairs [c] | OR (95% CI) for colorectal cancer risk relative to reference, [a] by percentile distribution of biomarkers [b] | | | | | *P* for overall association | *P* for nonlinear association | *P* for heterogeneity |
|---|---|---|---|---|---|---|---|---|---|
| | | 10th | 30th | 50th | 70th | 90th | | | |
| **Overall by anatomic site** | | | | | | | | | |
| 5-$F_{2t}$-IsoP | | | | | | | | | |
| Colon cancer | 1127 | 1.14 (1.03, 1.25) | 1.06 (1.02, 1.10) | Reference | 0.92 (0.86, 0.97) | 0.77 (0.61, 0.97) | 0.004 | 0.432 | 0.011 |
| Rectal cancer | 615 | 1.10 (0.96, 1.26) | 1.05 (0.99, 1.11) | Reference | 0.95 (0.90, 1.00) | 0.88 (0.76, 1.02) | 0.121 | 0.422 | |
| 15-$F_{2t}$-IsoP | | | | | | | | | |
| Colon cancer | 1031 | 1.00 (0.94, 1.07) | 1.00 (0.99, 1.02) | Reference | 0.98 (0.95, 1.01) | 0.91 (0.81, 1.02) | 0.287 | 0.618 | 0.312 |
| Rectal cancer | 586 | 0.97 (0.87, 1.07) | 0.99 (0.97, 1.02) | Reference | 1.01 (0.96, 1.05) | 0.97 (0.87, 1.08) | 0.575 | 0.400 | |
| 15-$F_{2t}$-IsoP-M | | | | | | | | | |
| Colon cancer | 1219 | 1.10 (1.02, 1.18) | 1.04 (1.00, 1.08) | Reference | 0.96 (0.90, 1.03) | 0.90 (0.74, 1.10) | 0.007 | 0.103 | 0.612 |
| Rectal cancer | 700 | 1.12 (1.04, 1.21) | 1.07 (1.02, 1.12) | Reference | 0.90 (0.82, 0.98) | 0.73 (0.55, 0.96) | 0.017 | 0.967 | |
| RPS-IsoP | | | | | | | | | |
| Colon cancer | 1023 | 1.28 (1.04, 1.56) | 1.11 (1.02, 1.21) | Reference | 0.92 (0.85, 0.98) | 0.87 (0.77, 0.98) | 0.059 | 0.018 | 0.481 |
| Rectal cancer | 604 | 1.09 (0.84, 1.41) | 1.03 (0.93, 1.15) | Reference | 0.97 (0.87, 1.07) | 0.96 (0.82, 1.12) | 0.548 | 0.501 | |
| **Colon cancer by time internal** | | | | | | | | | |
| 5-$F_{2t}$-IsoP | | | | | | | | | |
| Cases diagnosed <5 years | 228 | 1.67 (1.24, 2.25) | 1.25 (1.10, 1.41) | Reference | 0.79 (0.68, 0.91) | 0.64 (0.38, 1.08) | 0.001 | 0.024 | <0.001 |
| Cases diagnosed 5-9 years | 290 | 1.17 (0.95, 1.44) | 1.08 (0.98, 1.17) | Reference | 0.90 (0.82, 1.00) | 0.77 (0.54, 1.10) | 0.129 | 0.546 | |
| Cases diagnosed >9 years | 609 | 1.05 (0.95, 1.16) | 1.02 (0.97, 1.07) | Reference | 0.98 (0.88, 1.08) | 0.92 (0.64, 1.33) | 0.674 | 0.713 | |
| 15-$F_{2t}$-IsoP | | | | | | | | | |
| Cases diagnosed <5 years | 216 | 1.04 (0.91, 1.20) | 1.01 (0.97, 1.06) | Reference | 0.97 (0.89, 1.06) | 0.97 (0.81, 1.16) | 0.839 | 0.580 | 0.002 |
| Cases diagnosed 5-9 years | 222 | 0.96 (0.82, 1.13) | 1.00 (0.95, 1.04) | Reference | 0.95 (0.86, 1.05) | 0.71 (0.48, 1.04) | 0.199 | 0.228 | |
| Cases diagnosed >9 years | 593 | 1.01 (0.92, 1.10) | 1.00 (0.98, 1.03) | Reference | 0.98 (0.93, 1.03) | 0.92 (0.77, 1.10) | 0.657 | 0.825 | |
| 15-$F_{2t}$-IsoP-M | | | | | | | | | |
| Cases diagnosed <5 years | 243 | 1.14 (0.96, 1.34) | 1.02 (0.94, 1.11) | Reference | 1.02 (0.88, 1.19) | 1.07 (0.71, 1.63) | 0.031 | 0.021 | 0.231 |
| Cases diagnosed 5-9 years | 334 | 1.08 (0.95, 1.23) | 1.03 (0.96, 1.11) | Reference | 0.96 (0.86, 1.08) | 0.89 (0.65, 1.22) | 0.327 | 0.484 | |
| Cases diagnosed >9 years | 642 | 1.10 (0.97, 1.24) | 1.04 (0.97, 1.12) | Reference | 0.95 (0.83, 1.07) | 0.86 (0.61, 1.23) | 0.179 | 0.599 | |

| | | | | | | | | | |
|---|---|---|---|---|---|---|---|---|---|
| RPS-IsoP | | | | | | | | | |
| Cases diagnosed <5 years | 208 | 1.32 (0.82, 2.12) | 1.12 (0.92, 1.36) | Reference | 0.92 (0.79, 1.06) | 0.87 (0.69, 1.09) | 0.416 | 0.259 | 0.003 |
| Cases diagnosed 5-9 years | 222 | 1.50 (0.95, 2.37) | 1.19 (0.98, 1.43) | Reference | 0.88 (0.76, 1.02) | 0.82 (0.66, 1.04) | 0.185 | 0.075 | |
| Cases diagnosed >9 years | 593 | 1.23 (0.94, 1.60) | 1.10 (0.97, 1.24) | Reference | 0.92 (0.83, 1.03) | 0.89 (0.74, 1.07) | 0.245 | 0.108 | |
| **Rectal cancer by time internal** | | | | | | | | | |
| 5-$F_{2t}$-IsoP | | | | | | | | | |
| Cases diagnosed <5 years | 159 | 1.42 (0.98, 2.04) | 1.17 (1.01, 1.37) | Reference | 0.83 (0.70, 0.99) | 0.66 (0.36, 1.21) | 0.078 | 0.338 | 0.004 |
| Cases diagnosed 5-9 years | 179 | 1.10 (0.82, 1.47) | 1.04 (0.92, 1.18) | Reference | 0.95 (0.86, 1.06) | 0.91 (0.75, 1.09) | 0.577 | 0.670 | |
| Cases diagnosed >9 years | 313 | 1.06 (0.90, 1.25) | 1.03 (0.97, 1.11) | Reference | 0.94 (0.83, 1.05) | 0.82 (0.55, 1.20) | 0.519 | 0.923 | |
| 15-$F_{2t}$-IsoP | | | | | | | | | |
| Cases diagnosed <5 years | 151 | 1.19 (0.95, 1.47) | 1.04 (0.98, 1.11) | Reference | 0.96 (0.84, 1.09) | 1.15 (0.75, 1.78) | 0.213 | 0.079 | 0.034 |
| Cases diagnosed 5-9 years | 136 | 1.04 (0.84, 1.30) | 1.01 (0.95, 1.08) | Reference | 0.97 (0.86, 1.08) | 0.92 (0.78, 1.08) | 0.500 | 0.819 | |
| Cases diagnosed >9 years | 299 | 0.87 (0.76, 1.00) | 0.98 (0.95, 1.01) | Reference | 0.97 (0.89, 1.07) | 0.84 (0.58, 1.21) | 0.078 | 0.024 | |
| 15-$F_{2t}$-IsoP-M | | | | | | | | | |
| Cases diagnosed <5 years | 168 | 0.98 (0.76, 1.27) | 1.00 (0.85, 1.17) | Reference | 1.00 (0.75, 1.34) | 1.00 (0.42, 2.35) | 0.921 | 0.764 | 0.087 |
| Cases diagnosed 5-9 years | 210 | 1.19 (1.03, 1.38) | 1.09 (1.02, 1.16) | Reference | 0.91 (0.80, 1.04) | 0.78 (0.52, 1.15) | 0.053 | 0.363 | |
| Cases diagnosed >9 years | 327 | 1.17 (1.02, 1.34) | 1.10 (1.01, 1.19) | Reference | 0.85 (0.72, 1.00) | 0.61 (0.38, 1.00) | 0.072 | 0.649 | |
| RPS-IsoP | | | | | | | | | |
| Cases diagnosed <5 years | 141 | 0.72 (0.41, 1.25) | 0.88 (0.71, 1.09) | Reference | 1.13 (0.93, 1.37) | 1.22 (0.91, 1.63) | 0.197 | 0.263 | 0.011 |
| Cases diagnosed 5-9 years | 136 | 0.91 (0.53, 1.58) | 0.96 (0.77, 1.20) | Reference | 1.04 (0.84, 1.28) | 1.06 (0.77, 1.45) | 0.932 | 0.745 | |
| Cases diagnosed >9 years | 327 | 1.42 (0.97, 2.06) | 1.16 (0.99, 1.36) | Reference | 0.84 (0.70, 1.01) | 0.76 (0.56, 1.02) | 0.186 | 0.084 | |

Abbreviations: 5-$F_{2t}$-IsoP, 5-$F_{2t}$-Isoprostane; 15-$F_{2t}$-IsoP, 15-$F_{2t}$-Isoprostane; 15-$F_{2t}$-IsoP-M, 2,3-Dinor-5,6-dihydro-15-$F_{2t}$-Isoprostane; RPS-IsoP, ratio of 15-F2t-IsoP-M (metabolic product) to 15-F2t-IsoP (substrate); CI, confidence interval; ng/mg Cr, nanograms per milligram of creatinine; OR, odds ratio; SMHS, Shanghai Men's Health Study; SWHS, Shanghai Women's Health Study.

[a] Multivariable ORs estimated using a conditional logistic regression model with restricted cubic-spline functions, stratified by the time interval between the baseline sample collection and cancer diagnosis during follow-up, using the 50th percentile of biomarkers in controls as the reference; and multivariable adjusted for age at baseline, education, cigarette smoking, alcohol consumption, BMI, physical activity, regular use of vitamin supplements, regular use of aspirin and other nonsteroidal anti-inflammatory drugs, Charlson comorbidity score, family history of colorectal cancer in first-degree relatives, and total energy intake.

[b] Cutoffs were based on the percentile distribution of biomarker levels in controls in the SWHS and SMHS — for 5-$F_{2t}$-IsoP (ng/mg Cr): 1.15 (10th), 1.95 (30th), 2.79 (50th), 4.37 (70th), and 7.90 (90th) in the SWHS and SMHS; for 15-$F_{2t}$-IsoP (ng/mg Cr): 0.64 (10th), 1.00 (30th), 1.20 (50th), 1.91 (70th), and 3.6 (90th); for 15-$F_{2t}$-IsoP-M (ng/mg Cr): 4.30 (10th), 7.27 (30th), 10.70 (50th), 15.88 (70th), and 25.14 (90th); for RPS-IsoP: 272.6% (10th), 563.2% (30th), 810.3% (50th), 1140.9% (70th), and 1950.0% (90th).

[c] The number of case-control pairs after exclusion of batches with CV > 20%.

**Supplementary Table 9. Associations between baseline urinary levels of $F_2$-IsoPs and subsequent risk of colorectal cancer: analyses confined to batches with CV <20% and without inter-batch normalization**

| | Number of pairs [c] | OR (95% CI) for colorectal cancer risk relative to reference, [a] by percentile distribution of biomarkers [b] | | | | | *P* for overall association | *P* for nonlinear association |
|---|---|---|---|---|---|---|---|---|
| | | 10th | 30th | 50th | 70th | 90th | | |
| **In the SWHS and SMHS** | | | | | | | | |
| 5-$F_{2t}$-IsoP | 1778 | 1.18 (1.02, 1.38) | 1.08 (1.01, 1.16) | Reference | 0.90 (0.84, 0.96) | 0.79 (0.70, 0.90) | 0.001 | 0.190 |
| 15-$F_{2t}$-IsoP | 1617 | 0.94 (0.83, 1.05) | 0.98 (0.94, 1.02) | Reference | 1.04 (0.96, 1.13) | 1.00 (0.90, 1.12) | 0.174 | 0.182 |
| 15-$F_{2t}$-IsoP-M | 1919 | 1.20 (1.03- 1.39) | 1.10 (1.02- 1.18) | Reference | 0.90 (0.85- 0.95) | 0.78 (0.69- 0.89) | <0.001 | 0.330 |
| RPS-IsoP | 1599 | 1.22 (1.04, 1.43) | 1.09 (1.02, 1.16) | Reference | 0.93 (0.88, 0.99) | 0.90 (0.82, 0.98) | 0.044 | 0.014 |
| **In the SCCS** | | | | | | | | |
| 5-$F_{2t}$-IsoP | 285 | 1.37 (1.06, 1.76) | 1.20 (1.04, 1.39) | Reference | 0.84 (0.73, 0.98) | 0.85 (0.68, 1.06) | 0.030 | 0.010 |
| 15-$F_{2t}$-IsoP | 285 | 1.08 (0.84, 1.41) | 1.03 (0.94, 1.12) | Reference | 0.96 (0.84, 1.09) | 0.91 (0.73, 1.14) | 0.627 | 0.606 |
| 15-$F_{2t}$-IsoP-M | 285 | 1.25 (0.93- 1.67) | 1.10 (0.97- 1.25) | Reference | 0.93 (0.85- 1.03) | 0.95 (0.78- 1.14) | 0.329 | 0.140 |
| RPS-IsoP | 285 | 0.97 (0.69-1.35) | 0.99 (0.87-1.13) | Reference | 0.99 (0.90-1.08) | 0.93 (0.75-1.15) | 0.739 | 0.622 |

Abbreviations: : 5-$F_{2t}$-IsoP, 5-$F_{2t}$-Isoprostane; 15-$F_{2t}$-IsoP, 15-$F_{2t}$-Isoprostane; 15-$F_{2t}$-IsoP-M, 2,3-Dinor-5,6-dihydro-15-$F_{2t}$-Isoprostane; RPS-IsoP, ratio of 15-F2t-IsoP-M (metabolic product) to 15-F2t-IsoP (substrate); CI, confidence interval; ng/mg Cr, nanograms per milligram of creatinine; OR, odds ratio; SCCS, Southern Community Cohort Study; SMHS, Shanghai Men's Health Study; SWHS, Shanghai Women's Health Study.

[a] Multivariable ORs estimated using a conditional logistic regression model with restricted cubic-spline functions, using the 50th percentile of biomarkers in controls as the reference; and multivariable adjusted for age at baseline, education, cigarette smoking, alcohol consumption, BMI, physical activity, regular use of vitamin supplements, regular use of aspirin and other nonsteroidal anti-inflammatory drugs, Charlson comorbidity score, family history of colorectal cancer in first-degree relatives, and total energy intake.

[b] Cutoffs were based on the percentile distribution of biomarker levels in controls—for 5-$F_{2t}$-IsoP (ng/mg Cr): 1.15 (10th), 1.95 (30th), 2.79 (50th), 4.37 (70th), and 7.90 (90th) in the SWHS and SMHS; and 0.74 (10th), 1.18 (30th), 1.95 (50th), 3.08 (70th) and 6.01 (90th) in the SCCS; for 15-$F_{2t}$-IsoP (ng/mg Cr): 0.64 (10th), 1.00 (30th), 1.20 (50th), 1.91 (70th), and 3.6 (90th) in the SWHS and SMHS, and 0.54 (10th), 0.83 (30th), 1.08 (50th), 1.50 (70th) and 2.89 (90th) in the SCCS; for 15-$F_{2t}$-IsoP-M (ng/mg Cr): 4.30 (10th), 7.27 (30th), 10.70 (50th), 15.88 (70th), and 25.14 (90th) in the SWHS and SMHS; and 6.81 (10th), 10.49 (30th), 14.75 (50th), 20.34 (70th) and 32.41 (90th) in the SCCS; for RPS-IsoP: 272.6% (10th), 563.2% (30th), 810.3% (50th), 1140.9% (70th), and 1950.0% (90th) in the SWHS and SMHS, and 553.0% (10th), 1003.3% (30th), 1362.1% (50th), 1835.6% (70th) and 2586.6% (90th) in the SCCS.

[c] The number of case-control pairs after exclusion of batches with CV >20%, with the case-control ratio of 1:1 in the SWHS, 1:1 in the SMHS, and 1:2 in the SCCS.

**Supplementary Table 10. Associations between baseline urinary levels of $F_2$-IsoPs and subsequent risk of colorectal cancer by follow-up time: analyses confined to batches with CV <20% and without inter-batch normalization in the SWHS and SMHS**

| | Number of pairs [c] | OR (95% CI) for colorectal cancer risk relative to reference, [a] by percentile distribution of biomarkers [b] | | | | | *P* for overall association | *P* for nonlinear association |
|---|---|---|---|---|---|---|---|---|
| | | 10th | 30th | 50th | 70th | 90th | | |
| **Cases diagnosed <5 years after enrollment** | | | | | | | | |
| 5-$F_{2t}$-IsoP | 387 | 2.01 (1.38- 2.93) | 1.41 (1.17- 1.68) | Reference | 0.63 (0.51- 0.78) | 0.43 (0.30- 0.62) | <0.001 | 0.009 |
| 15-$F_{2t}$-IsoP | 367 | 1.16 (0.94- 1.42) | 1.05 (0.98- 1.13) | Reference | 0.88 (0.74- 1.05) | 0.83 (0.63- 1.10) | 0.388 | 0.180 |
| 15-$F_{2t}$-IsoP-M | 411 | 1.16 (0.86- 1.57) | 1.08 (0.94- 1.24) | Reference | 0.92 (0.81- 1.04) | 0.82 (0.61- 1.09) | 0.352 | 0.654 |
| RPS-IsoP | 349 | 1.04 (0.73- 1.47) | 1.01 (0.88- 1.16) | Reference | 0.99 (0.89- 1.11) | 0.99 (0.84- 1.17) | 0.731 | 0.829 |
| **Cases diagnosed 5-9 years after enrollment** | | | | | | | | |
| 5-$F_{2t}$-IsoP | 469 | 1.20 (0.92- 1.55) | 1.09 (0.96- 1.24) | Reference | 0.89 (0.77- 1.03) | 0.81 (0.65- 1.01) | 0.169 | 0.284 |
| 15-$F_{2t}$-IsoP | 358 | 0.95 (0.75- 1.21) | 0.98 (0.91- 1.06) | Reference | 1.03 (0.86- 1.24) | 1.00 (0.76- 1.31) | 0.504 | 0.572 |
| 15-$F_{2t}$-IsoP-M | 539 | 1.22 (0.92- 1.62) | 1.11 (0.97- 1.26) | Reference | 0.90 (0.82- 0.99) | 0.80 (0.66- 0.96) | 0.051 | 0.461 |
| RPS-IsoP | 358 | 1.27 (0.90- 1.78) | 1.10 (0.96- 1.27) | Reference | 0.92 (0.82- 1.04) | 0.89 (0.74- 1.06) | 0.382 | 0.171 |
| **Cases diagnosed >9 years after enrollment** | | | | | | | | |
| 5-$F_{2t}$-IsoP | 922 | 1.04 (0.83- 1.31) | 1.02 (0.93, 1.13) | Reference | 0.96 (0.88, 1.04) | 0.87 (0.70, 1.08) | 0.436 | 0.983 |
| 15-$F_{2t}$-IsoP | 892 | 0.83 (0.69- 1.01) | 0.95 (0.89- 1.00) | Reference | 1.09 (0.98- 1.20) | 1.01 (0.86- 1.19) | 0.123 | 0.045 |
| 15-$F_{2t}$-IsoP-M | 969 | 1.21 (0.96- 1.52) | 1.10 (0.99- 1.24) | Reference | 0.88 (0.81- 0.97) | 0.75 (0.62- 0.91) | 0.011 | 0.575 |
| RPS-IsoP | 892 | 1.31 (1.05- 1.62) | 1.12 (1.02- 1.24) | Reference | 0.89 (0.81- 0.98) | 0.84 (0.72- 0.98) | 0.046 | 0.013 |

Abbreviations: : 5-$F_{2t}$-IsoP, 5-$F_{2t}$-Isoprostane; 15-$F_{2t}$-IsoP, 15-$F_{2t}$-Isoprostane; 15-$F_{2t}$-IsoP-M, 2,3-Dinor-5,6-dihydro-15-$F_{2t}$-Isoprostane; RPS-IsoP, ratio of 15-F2t-IsoP-M (metabolic product) to 15-F2t-IsoP (substrate); CI, confidence interval; ng/mg Cr, nanograms per milligram of creatinine; OR, odds ratio; SMHS, Shanghai Men's Health Study; SWHS, Shanghai Women's Health Study.

[a] Multivariable ORs estimated using a conditional logistic regression model with restricted cubic-spline functions, stratified by the time interval between the baseline sample collection and cancer diagnosis during follow-up, using the 50th percentile of biomarkers in controls as the reference; and multivariable adjusted for age at baseline, education, cigarette smoking, alcohol consumption, BMI, physical activity, regular use of vitamin supplements, regular use of aspirin and other nonsteroidal anti-inflammatory drugs, Charlson comorbidity score, family history of colorectal cancer in first-degree relatives, and total energy intake.

[b] Cutoffs were based on the percentile distribution of biomarker levels in controls in the SWHS and SMHS — for 5-$F_{2t}$-IsoP (ng/mg Cr): 1.15 (10th), 1.95 (30th), 2.79 (50th), 4.37 (70th), and 7.90 (90th); for 15-$F_{2t}$-IsoP (ng/mg Cr): 0.64 (10th), 1.00 (30th), 1.20 (50th), 1.91 (70th), and 3.6 (90th); for 15-$F_{2t}$-IsoP-M (ng/mg Cr): 4.30 (10th), 7.27 (30th), 10.70 (50th), 15.88 (70th), and 25.14 (90th); for RPS-IsoP: 272.6% (10th), 563.2% (30th), 810.3% (50th), 1140.9% (70th), and 1950.0% (90th).

[c] The number of case-control pairs after exclusion of batches with CV >20%.

**Supplementary Table 11. Associations between baseline urinary levels of F2-IsoPs and subsequent risk of colorectal cancer: analyses in all participants without inter-batch normalization**

| | OR (95% CI) for colorectal cancer risk relative to reference, [a] by percentile distribution of biomarkers [b] | | | | | *P* for overall association | *P* for nonlinear association |
|---|---|---|---|---|---|---|---|
| | **10th** | **30th** | **50th** | **70th** | **90th** | | |
| **In the SWHS and SMHS** | | | | | | | |
| 5-$F_{2t}$-IsoP | 1.19 (1.04, 1.36) | 1.09 (1.02, 1.16) | Reference | 0.90 (0.84, 0.96) | 0.80 (0.71, 0.90) | 0.001 | 0.108 |
| 15-$F_{2t}$-IsoP | 1.00 (0.91, 1.10) | 1.00 (0.97, 1.03) | Reference | 0.99 (0.93, 1.06) | 0.95 (0.86, 1.05) | 0.194 | 0.758 |
| 15-$F_{2t}$-IsoP-M | 1.25 (1.06, 1.47) | 1.12 (1.03, 1.21) | Reference | 0.90 (0.85, 0.95) | 0.79 (0.70, 0.89) | 0.000 | 0.197 |
| RPS-IsoP | 1.15 (1.02, 1.30) | 1.07 (1.01, 1.13) | Reference | 0.95 (0.90, 0.99) | 0.92 (0.85, 0.99) | 0.060 | 0.022 |
| **In the SCCS** | | | | | | | |
| 5-$F_{2t}$-IsoP | 1.37 (1.06, 1.76) | 1.20 (1.04, 1.39) | Reference | 0.84 (0.73, 0.98) | 0.85 (0.68, 1.06) | 0.03 | 0.01 |
| 15-$F_{2t}$-IsoP | 1.08 (0.84, 1.41) | 1.03 (0.94, 1.12) | Reference | 0.96 (0.84, 1.09) | 0.91 (0.73, 1.14) | 0.627 | 0.606 |
| 15-$F_{2t}$-IsoP-M | 1.25 (0.93, 1.67) | 1.10 (0.97, 1.25) | Reference | 0.93 (0.85, 1.03) | 0.95 (0.78, 1.14) | 0.329 | 0.14 |
| RPS-IsoP | 0.97 (0.69, 1.35) | 0.99 (0.87, 1.13) | Reference | 0.99 (0.90, 1.08) | 0.93 (0.75, 1.15) | 0.739 | 0.622 |

Abbreviations: 5-$F_{2t}$-IsoP, 5-$F_{2t}$-Isoprostane; 15-$F_{2t}$-IsoP, 15-$F_{2t}$-Isoprostane; 15-$F_{2t}$-IsoP-M, 2,3-Dinor-5,6-dihydro-15-$F_{2t}$-Isoprostane; RPS-IsoP, ratio of 15-F2t-IsoP-M (metabolic product) to 15-F2t-IsoP (substrate); CI, confidence interval; ng/mg Cr, nanograms per milligram of creatinine; OR, odds ratio; SCCS, Southern Community Cohort Study; SMHS, Shanghai Men's Health Study; SWHS, Shanghai Women's Health Study.

[a] The OR was estimated using a conditional logistic regression model with restricted cubic-spline functions, with the 50th percentile of biomarkers in controls treated as the reference, multivariable adjusted for age at baseline, education, cigarette smoking, alcohol consumption, BMI, physical activity, regular use of vitamin supplements, regular use of aspirin and other nonsteroidal anti-inflammatory drugs, Charlson comorbidity score, family history of colorectal cancer in first-degree relatives, and total energy intake. The number of case-control pairs included in the analysis without batch-CV-based exclusion was 1938 in the SWHS and SMHS and 285 in the SCCS, with the case-control ratio of 1:1 in the SWHS and SMHS, and 1:2 in the SCCS.

[b] Cutoffs were based on the percentile distribution of biomarker levels in controls—for 5-$F_{2t}$-IsoP (ng/mg Cr): 1.15 (10th), 1.95 (30th), 2.79 (50th), 4.37 (70th), and 7.90 (90th) in the SWHS and SMHS; and 0.74 (10th), 1.18 (30th), 1.95 (50th), 3.08 (70th) and 6.01 (90th) in the SCCS; for 15-$F_{2t}$-IsoP (ng/mg Cr): 0.64 (10th), 1.00 (30th), 1.20 (50th), 1.91 (70th), and 3.6 (90th) in the SWHS and SMHS, and 0.54 (10th), 0.83 (30th), 1.08 (50th), 1.50 (70th) and 2.89 (90th) in the SCCS. for 15-$F_{2t}$-IsoP-M (ng/mg Cr): 4.30 (10th), 7.27 (30th), 10.70 (50th), 15.88 (70th), and 25.14 (90th) in the SWHS and SMHS; and 6.81 (10th), 10.49 (30th), 14.75 (50th), 20.34 (70th) and 32.41 (90th) in the SCCS; for RPS-IsoP: 272.6% (10th), 563.2% (30th), 810.3% (50th), 1140.9% (70th), and 1950.0% (90th) in the SWHS and SMHS, and 553.0% (10th), 1003.3% (30th), 1362.1% (50th), 1835.6% (70th) and 2586.6% (90th) in the SCCS.

**Supplementary Table 12. Associations between baseline urinary levels of F2-IsoPs and subsequent risk of colorectal cancer stratified by follow-up time: analyses in all participants from the SWHS and SMHS and without inter-batch normalization**

| | OR (95% CI) for colorectal cancer risk relative to reference,[s] by percentile distribution of biomarkers [b] | | | | | *P* for overall association | *P* for nonlinear association |
|---|---|---|---|---|---|---|---|
| | 10th | 30th | 50th | 70th | 90th | | |
| **Cases diagnosed <5 years after enrollment** | | | | | | | |
| 5-$F_{2t}$-IsoP | 2.09 (1.39, 3.13) | 1.52 (1.21, 1.90) | Reference | 0.74 (0.65, 0.85) | 0.62 (0.46, 0.83) | 0.000 | 0.011 |
| 15-$F_{2t}$-IsoP | 1.21 (0.94, 1.55) | 1.08 (0.98, 1.19) | Reference | 0.91 (0.80, 1.03) | 0.89 (0.73, 1.08) | 0.335 | 0.152 |
| 15-$F_{2t}$-IsoP-M | 1.43 (0.99, 2.06) | 1.20 (1.00, 1.46) | Reference | 0.85 (0.73, 0.99) | 0.76 (0.55, 1.05) | 0.116 | 0.140 |
| RPS-IsoP | 1.02 (0.81, 1.28) | 1.01 (0.92, 1.11) | Reference | 0.99 (0.88, 1.12) | 0.98 (0.71, 1.35) | 0.969 | 0.879 |
| **Cases diagnosed 5-9 years after enrollment** | | | | | | | |
| 5-$F_{2t}$-IsoP | 1.32 (1.00, 1.75) | 1.16 (1.00, 1.34) | Reference | 0.87 (0.76, 0.98) | 0.79 (0.66, 0.96) | 0.050 | 0.116 |
| 15-$F_{2t}$-IsoP | 1.01 (0.83, 1.22) | 1.00 (0.92, 1.10) | Reference | 0.98 (0.85, 1.12) | 0.89 (0.73, 1.08) | 0.230 | 0.806 |
| 15-$F_{2t}$-IsoP-M | 1.25 (0.90, 1.75) | 1.11 (0.96, 1.28) | Reference | 0.91 (0.84, 1.00) | 0.81 (0.67, 0.96) | 0.053 | 0.464 |
| RPS-IsoP | 1.09 (0.88, 1.33) | 1.03 (0.95, 1.13) | Reference | 0.96 (0.87, 1.06) | 0.95 (0.82, 1.09) | 0.680 | 0.433 |
| **Cases diagnosed >9 years after enrollment** | | | | | | | |
| 5-$F_{2t}$-IsoP | 1.01 (0.83, 1.23) | 1.01 (0.92, 1.10) | Reference | 0.99 (0.91, 1.06) | 0.91 (0.77, 1.07) | 0.439 | 0.789 |
| 15-$F_{2t}$-IsoP | 0.92 (0.79, 1.06) | 0.98 (0.95, 1.01) | Reference | 1.05 (0.96, 1.16) | 1.04 (0.90, 1.20) | 0.361 | 0.201 |
| 15-$F_{2t}$-IsoP-M | 1.20 (0.94, 1.53) | 1.10 (0.98, 1.25) | Reference | 0.90 (0.83, 0.97) | 0.77 (0.65, 0.93) | 0.012 | 0.752 |
| RPS-IsoP | 1.29 (1.08, 1.55) | 1.12 (1.03, 1.22) | Reference | 0.93 (0.88, 0.98) | 0.90 (0.83, 0.98) | 0.020 | 0.006 |

Abbreviations: 5-$F_{2t}$-IsoP, 5-$F_{2t}$-Isoprostane; 15-$F_{2t}$-IsoP, 15-$F_{2t}$-Isoprostane; 15-$F_{2t}$-IsoP-M, 2,3-Dinor-5,6-dihydro-15-$F_{2t}$-Isoprostane; RPS-IsoP, ratio of 15-$F_{2t}$-IsoP-M (β-oxidation product) to 15-$F_{2t}$-IsoP (substrate); CI, confidence interval; ng/mg Cr, nanograms per milligram of creatinine; OR, odds ratio; SMHS, Shanghai Men's Health Study; SWHS, Shanghai Women's Health Study.

[a] The OR was estimated using a conditional logistic regression model with restricted cubic-spline functions, with the 50th percentile of biomarkers in controls treated as the reference, multivariable adjusted for age at baseline, education, cigarette smoking, alcohol consumption, BMI, physical activity, regular use of vitamin supplements, regular use of aspirin and other nonsteroidal anti-inflammatory drugs, Charlson comorbidity score, family history of colorectal cancer in first-degree relatives, and total energy intake. The number of case-control pairs without batch-CV-based exclusion in SWHS and SMHS was 429 pairs in the cases diagnosed <5 years, 540 pairs in the cases diagnosed 5-9 years, and 969 pairs in the cases diagnosed >9 years, with the case-control ratio of 1:1.

[b] Cutoffs were based on the percentile distribution of biomarker levels in controls in the SWHS and SMHS — for 5-$F_{2t}$-IsoP (ng/mg Cr): 1.15 (10th), 1.95 (30th), 2.79 (50th), 4.37 (70th), and 7.90 (90th); for 15-$F_{2t}$-IsoP (ng/mg Cr): 0.64 (10th), 1.00 (30th), 1.20 (50th), 1.91 (70th), and 3.6 (90th); for 15-$F_{2t}$-IsoP-M (ng/mg Cr): 4.30 (10th), 7.27 (30th), 10.70 (50th), 15.88 (70th), and 25.14 (90th); for RPS-IsoP: 272.6% (10th), 563.2% (30th), 810.3% (50th), 1140.9% (70th), and 1950.0% (90th).

#### Supplementary Table 13. Associations between the index of DNA, RNA, and lipid OxS markers and subsequent risk of colorectal cancer

| | Number of pairs [c] | OR (95% CI) for colorectal cancer risk relative to reference, [a] by percentile distribution of the index [b] | | | | | *P* for overall association | *P* for nonlinear association |
|---|---|---|---|---|---|---|---|---|
| | | 10th | 30th | 50th | 70th | 90th | | |
| **In the SWHS and SMHS** | | | | | | | | |
| Overall [d] | 1778 | 1.28 (1.09, 1.51) | 1.12 (1.04, 1.20) | Reference | 0.88 (0.83, 0.94) | 0.77 (0.67, 0.87) | <0.001 | 0.088 |
| Cases diagnosed <5 years [d,e] | 387 | 2.11 (1.44, 3.09) | 1.43 (1.20, 1.70) | Reference | 0.62 (0.52, 0.75) | 0.36 (0.25, 0.51) | <0.001 | 0.062 |
| Cases diagnosed 5-9 years [d,e] | 469 | 1.33 (0.98, 1.81) | 1.14 (0.99, 1.32) | Reference | 0.86 (0.74, 1.00) | 0.80 (0.64, 1.00) | 0.146 | 0.111 |
| Cases diagnosed >9 years [d,e] | 922 | 1.07 (0.85, 1.35) | 1.03 (0.94, 1.13) | Reference | 0.95 (0.89, 1.02) | 0.86 (0.69, 1.06) | 0.317 | 0.993 |
| **In the SCCS** | | | | | | | | |
| Overall [d,e] | 285 | 1.69 (1.24, 2.30) | 1.21 (1.08, 1.35) | Reference | 0.86 (0.78, 0.95) | 0.89 (0.75, 1.06) | 0.004 | 0.001 |

Abbreviations: 5-$F_{2t}$-IsoP, 5-$F_{2t}$-Isoprostane; 8-oxo-dG, 8-oxo-7,8-dihydro-2'-deoxyguanosine; 8-oxo-Guo, 7,8-dihydro-8-oxo-guanosine; CI, confidence interval; ng/mg Cr, nanograms per milligram of creatinine; OR, odds ratio; SCCS, Southern Community Cohort Study; SMHS, Shanghai Men's Health Study; SWHS, Shanghai Women's Health Study.

[a] The OR was estimated by using a conditional logistic regression model with restricted cubic-spline functions, with the 50th percentile of the index in controls treated as the reference.

[b] The index is derived from the first principal component (PC1) from a principal component analysis (PCA) of three OxS markers: 5-$F_{2t}$-IsoP, 8-oxo-dG and 8-oxo-Guo. Cutoffs were based on the percentile distribution of the index levels in controls— -1.03 (10th), -0.67 (30th), -0.26 (50th), 0.290 (70th), and 1.54 (90th) in the SWHS and the SMHS; and –1.03 (10th), -0.69 (30th), -0.27 (50th), 0.22 (70th) and 1.60 (90th) in the SCCS.

[c] The number of case-control pairs after exclusion of batches with CV >20%, with case-control ratio of 1:1 in the SWHS, 1:1 in the SMHS and 1:2 in the SCCS.

[d] Multivariable adjusted for age at baseline, education, cigarette smoking, alcohol consumption, BMI, physical activity, regular use of vitamin supplements, regular use of aspirin and other nonsteroidal anti-inflammatory drugs, Charlson comorbidity score, family history of colorectal cancer in first-degree relatives, and total energy intake.

[e] Time interval between urine sample collection at baseline and cancer diagnosis during follow-up in the SWHS and SMHS was used to stratify the SWHS and SMHS, with *P* for heterogeneity across time internal groups < 0.001, while not in the SCCS as number of pairs was insufficient for the stratified analysis.

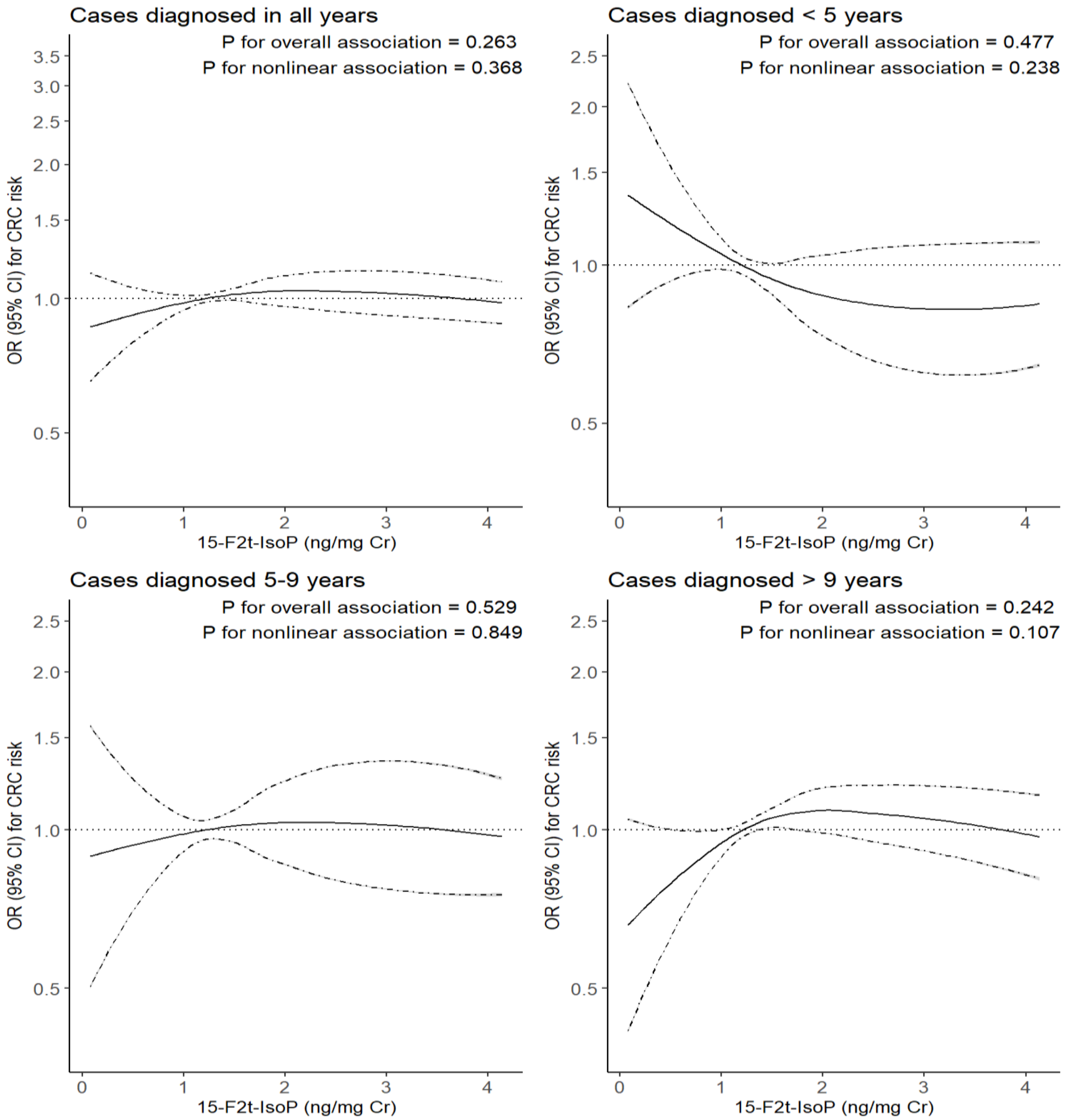


**Supplementary Figure 1**

**Figure Legend**

**Supplementary Figure 1. Smoothed plot for multivariable-adjusted odds ratios for colorectal cancer risk according to urinary levels of 15-$F_{2t}$-IsoP in the SWHS and the SMHS, by time interval from baseline to cancer diagnosis: Overall, <5 years, 5-9 years and >9 years.**

The OR was estimated using a conditional logistic regression model with restricted cubic spline functions, with the 50th percentile of biomarker levels in controls as the reference and adjusted for potential confounding factors as listed in the footnotes of Table 1. OR values on the *y* axis are shown in log scale. The solid line indicates the point estimate, and the dashed lines indicate the 95% CI.

Abbreviations: 15-$F_{2t}$-IsoP, 15-$F_{2t}$-Isoprostane; CI, confidence interval; ng/mg Cr, nanograms per milligram of creatinine; OR, odds ratio; SMHS, Shanghai Men's Health Study; SWHS, Shanghai Women's Health Study.

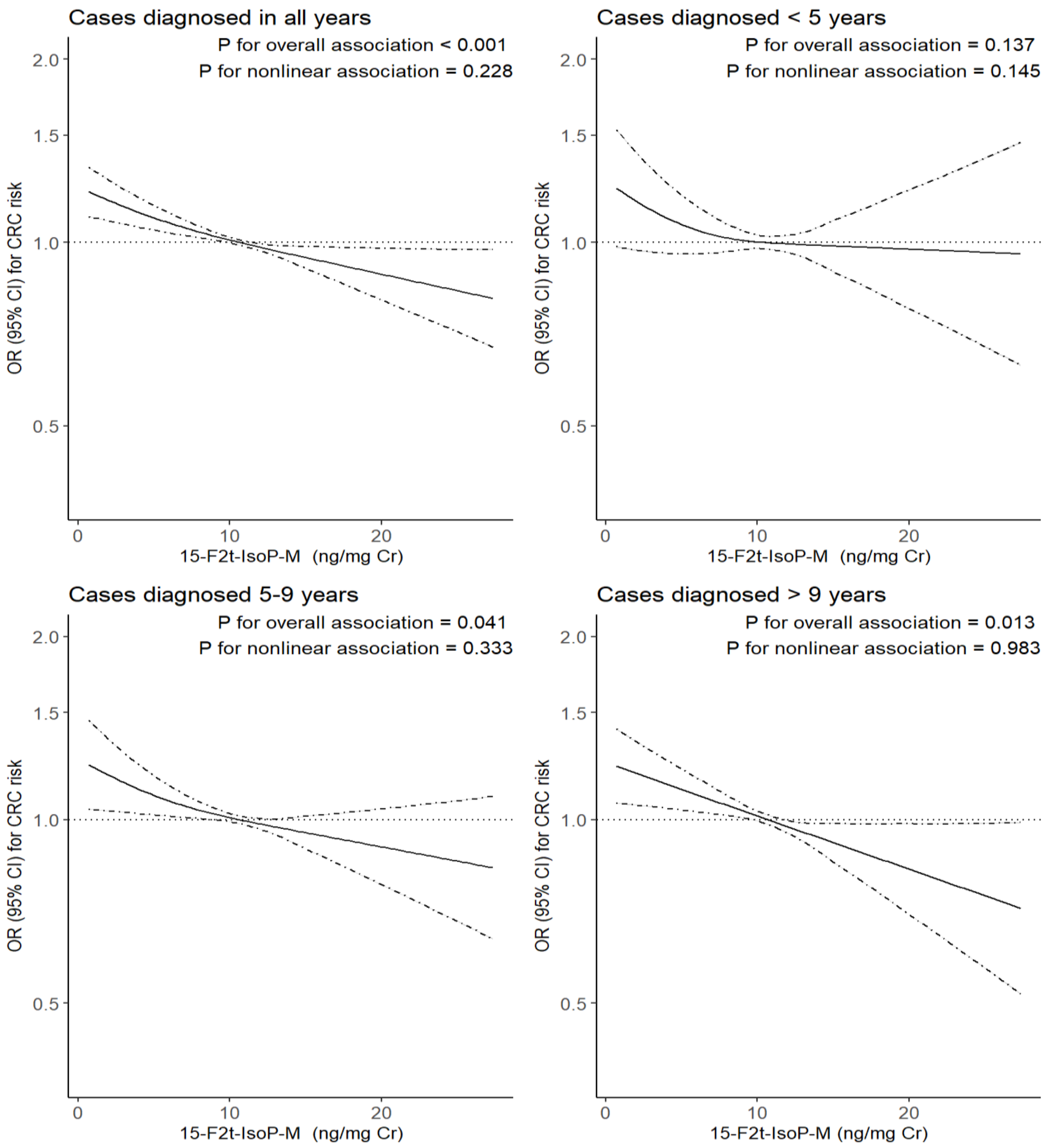


**Supplementary Figure 2**

**Figure Legend**

**Supplementary Figure 2. Smoothed plot for multivariable-adjusted odds ratios for colorectal cancer risk according to urinary levels of 15-$F_{2t}$-IsoP-M in the SWHS and the SMHS, by time interval from baseline to cancer diagnosis: Overall, <5 years, 5-9 years and >9 years.**

The OR was estimated using a conditional logistic regression model with restricted cubic spline functions, with the 50th percentile of biomarker levels in controls as the reference and adjusted for potential confounding factors as listed in the footnotes of Table 1. OR values on the *y* axis are shown in log scale. The solid line indicates the point estimate, and the dashed lines indicate the 95% CI.

Abbreviations: 15-$F_{2t}$-IsoP-M, 2,3-Dinor-5,6-dihydro-15-$F_{2t}$-Isoprostane; CI, confidence interval; ng/mg Cr, nanograms per milligram of creatinine; OR, odds ratio; SMHS, Shanghai Men's Health Study; SWHS, Shanghai Women's Health Study.

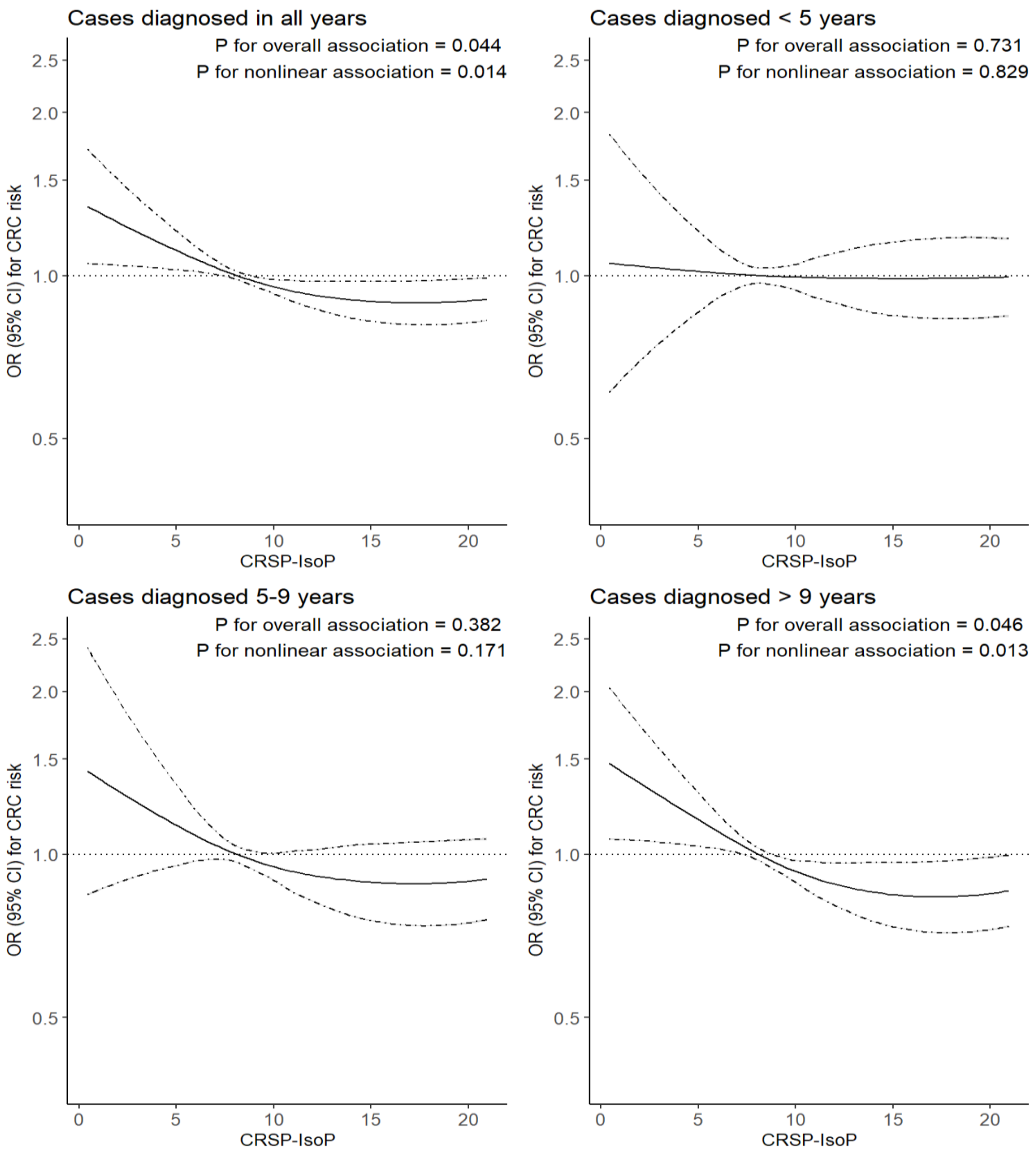


**Supplementary Figure 3**

**Figure Legend**

**Supplementary Figure 3. Smoothed plot for multivariable-adjusted odds ratios for colorectal cancer risk according to urinary levels of RPS-IsoP in the SWHS and the SMHS, by time interval from baseline to cancer diagnosis: Overall, <5 years, 5-9 years and >9 years.**

The OR was estimated using a conditional logistic regression model with restricted cubic spline functions, with the 50th percentile of biomarker levels in controls as the reference and adjusted for potential confounding factors as listed in the footnotes of Table 1. OR values on the *y* axis are shown in log scale. The solid line indicates the point estimate, and the dashed lines indicate the 95% CI.

Abbreviations: RPS-IsoP, ratio of 15-$F_{2t}$-IsoP-M (β-oxidation product) to 15-$F_{2t}$-IsoP (substrate); CI, confidence interval; ng/mg Cr, nanograms per milligram of creatinine; OR, odds ratio; SMHS, Shanghai Men's Health Study; SWHS, Shanghai Women's Health Study.

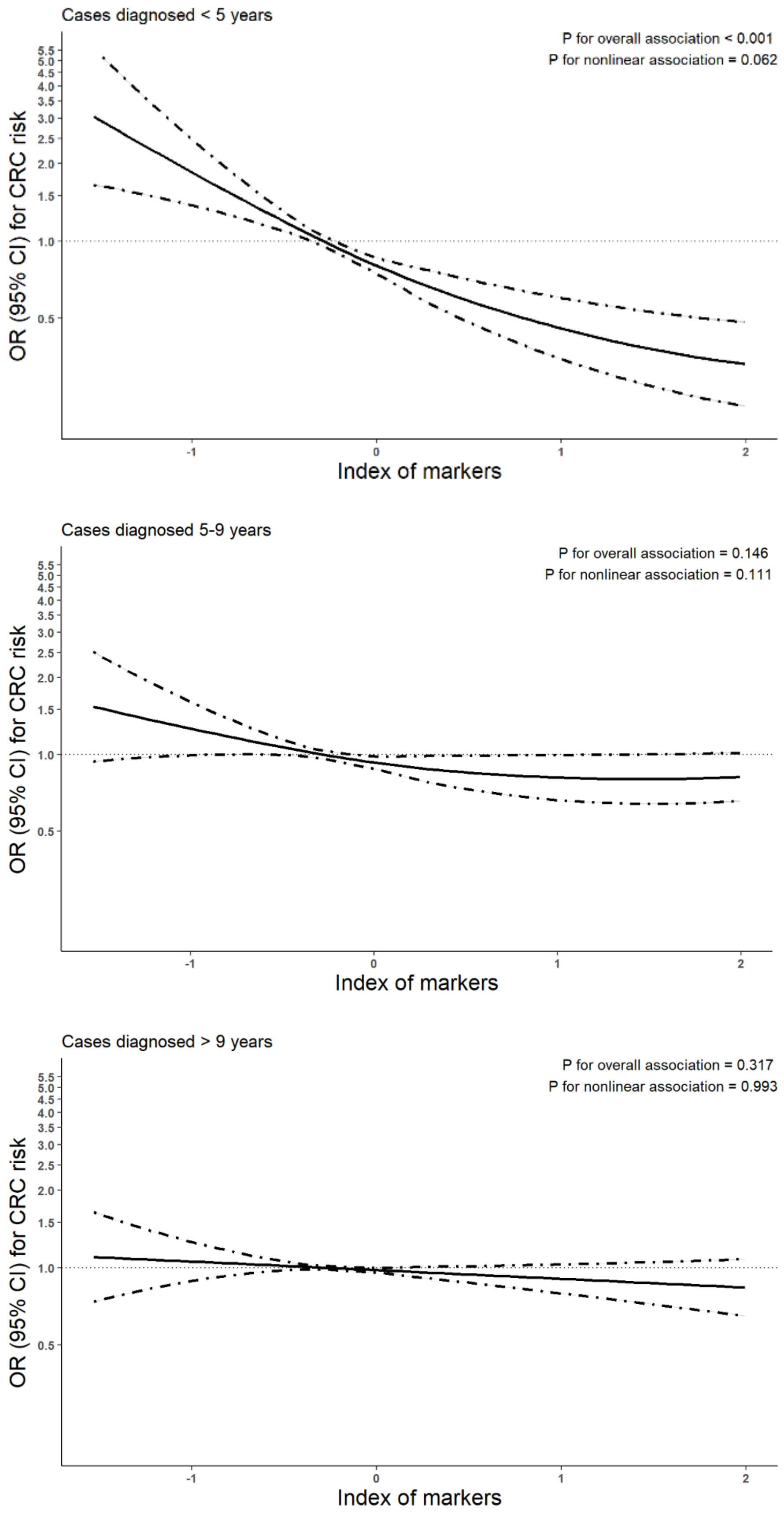


**Supplementary Figure 4**

**Figure Legend**

**Supplementary Figure 4. Smoothed plot for multivariable-adjusted ORs for colorectal cancer risk according to OxS marker index in the SWHS and SMHS, by time interval from baseline to cancer diagnosis: Overall, <5 years, 5-9 years and >9 years.**

The OR was estimated using a conditional logistic regression model with restricted cubic spline functions, with the 50th percentile of the OxS index in controls as the reference and adjusted for potential confounding factors as listed in the footnotes of Table 1. The index was constructed using the principal component analysis for 8-oxo-dG (DNA), 8-oxo-Guo (RNA) and 5-$F_{2t}$-IsoP (lipid) OxS markers. OR values on the *y* axis are shown in log scale. The solid line indicates the point estimate, and the dashed lines indicate the 95% CI.

Abbreviations: 5-$F_{2t}$-IsoP, 5-$F_{2t}$-Isoprostane; 8-oxo-dG, 8-oxo-7,8-dihydro-2'-deoxyguanosine; 8-oxo-Guo, 7,8-dihydro-8-oxo-guanosine; CI, confidence interval; ng/mg Cr, nanograms per milligram of creatinine; OR, odds ratio; SMHS, Shanghai Men's Health Study; SWHS, Shanghai Women's Health Study.
